# Rare variants in infection response protein pathways associated with sepsis in children

**DOI:** 10.1101/2025.06.12.25329504

**Authors:** Dylan Lawless, Pauline Rogg, Manon Bouzereau, Robin Fallegger, Zaira Seferbekova, Valeriia Timonina, Konstantin Popadin, Ali Saadat, Zhi Ming Xu, Simon Boutry, Christian W. Thorball, Flavia Hodel, Alessandro Borghesi, Johannes Trück, Martin Stocker, Klara M Posfay-Barbe, Ulrich Heininger, Sara Bernhard-Stirnemann, Anita Niederer-Loher, Christian Kahlert, Giancarlo Natalucci, Christa Relly, Thomas Riedel, Christoph Aebi, Christoph Berger, Eric Giannoni, Philipp Agyeman, Jacques Fellay, Luregn J Schlapbach, the Swiss Pediatric Sepsis Study

**Author notes:** These authors contributed equally to the work.

## Abstract

Sepsis is a major contributor to mortality in paediatric patients. We investigated the human genetic underpinnings of sepsis by analysing a large paediatric cohort with blood-culture confirmed sepsis through multi-tiered genomic assessments: single-case, single-variant, single-gene, and protein pathway analyses. We designed and applied novel analytical methods to automate for unbiased interpretation. We identified two pathways involved in susceptibility to sepsis, which contained 50 genes (42 and 8, respectively) including *KIR2DL4*, *KIR3DL3*, *KLRD1*, *LILRA1*, *SIGLEC1*, and *SIRPG*. These pathways are central to immune cell regulation, antigen processing, cellular signalling, and prevention of excessive inflammatory responses. A third enriched pathway of 22 genes was related to regulation of transcription. We additionally found 66 variants for inborn errors of immunity. Our findings highlight the influence of deleterious genomic variants on a shared immunological phenotype resulting in sepsis vulnerability in children. These insights lay a foundation for more personalised approaches to sepsis in children.

## 2 Introduction

Sepsis is a critical condition triggered by a dysregulated host response to infection causing tissue and organ damage. It is a major global public health concern and especially lethal in children (1). While some studies have pinpointed common human genetic variants linked to sepsis susceptibility, the role of rare variants remains underexplored (2),(3). The host genetic component of severe infectious diseases often eludes detection due to genetic heterogeneity (4). A shared immunological phenotype might be observed even when patients have unique variants that affect separate genes. This can occur when these genes code for proteins within the same functional pathway and thus result in similar phenotypic consequences. Genome-wide association studies (GWAS) assess individual variants. For common variants that are shared among patients, an association with disease can be relatively easy to detect. In rare disease and rare variant cohorts, GWAS may be underpowered even if there is an ostensibly shared phenotype among patients. In cohorts with rare variants, variant set association testing (VSAT) is common practice. This shift from a single variant level to gene-level and pathway-level analyses significantly increases complexity. Furthermore, a key limitation in similar studies is the use of inappropriate statistical tests and the bias of selectively favouring certain gene sets.

Our approach comprised four distinct stages to address these issues. First, we searched for potentially causal variants in single cases, adhering to the traditional genetic approach. Second, we conducted single variant association tests using both cases and controls. Third, we performed gene-level association tests, based on the collapse of common and rare variants. Finally, we performed pathway-level association analyses using precalculated protein interactions to ensure comprehensive and unbiased assessments. In all four stages, the current state of the art for the interpretation and reporting process relied on key reference databases (5),(6),(7),(8),(9),(10),(11),(12), which allowed us to define qualifying variants (QV) (13),(14), as well as stringent standards to interpret disease-associated variants (15),(16),(17). This approach represents a shift towards integrating gene-level and pathway-level analyses to tackle the challenges posed by rare variants in sepsis susceptibility, leveraging advanced statistical methodologies to uncover the genetic underpinnings of this complex condition. Using exome sequencing data from a large population-based cohort of children with blood culture-confirmed sepsis, part of which was previously investigated (18)(19), we employed optimised sequence kernel association tests (SKAT-O) for both rare and common variants in mixed VSAT (20),(21),(22). These methods, thoroughly reviewed (23) and previously successful in other diseases (24)(25), are complemented by innovations in protein pathway clustering (11) to enable us to discover enriched protein-coding rare genetic variants within shared pathways.

## 3 Results

### 3.1 Study participants

Our cohort consisted of 940 individuals: 534 Swiss children with blood culture-proven sepsis (26), 14 parents, and 392 in-house controls from the Swiss population. The control cohort was 76.79% adult male (301 of 392), therefore our analysis included statistical control for age and sex and was restricted to germline genomic variation. The cases consisted of blood-culture positive infections, of whom 325 (60.86%) were male and 209 (39.14%) were female. The age distribution was as follows: 91 (17.04%) were preterm neonates, 67 (12.55%) were term neonates, 94 (17.60%) were infants (1 month-1 year), 123 (23.03%) were toddlers (1-4 years), 79 (14.79%) were children (5-9 years), and 80 (14.98%) were children (10-16 years). A total of 251 individuals (47.99%) required intensive care unit (ICU) admission, and comorbidities were present in 274 individuals (52.39%). Hospital stay following blood culture sampling had a median duration of 16 days (range: 0-803, SD: 58.26), while PICU stay had a median duration of 11 days (range: −2-216, SD: 40.01). The most predominant pathogen groups in descending order were *Staphylococcus aureus* (18.06%), *Escherichia coli* (17.67%), *Streptococcus pneumoniae* (12.43%), *coagulase-negative staphylococci* (11.84%), other Gram-negative bacteria (8.54%), Group A and B streptococci (6.60 - 4.85%). Other pathogens each accounted for 2-5% of cases - viridans group streptococci, *Klebsiella spp*., *Enterococcus spp*., Haemophilus influenzae, *Pseudomonas aeruginosa*, fungal pathogens, and other Gram-positive bacteria. Hospital acquired infection was detected for 186 patients (35.57%). Additional features are reported in Table 1.

### 3.2 Single-case analysis

A concise overview of the analysis pipeline is depicted in **Figure 1 (and Figure S1).** Single-case analysis was performed using a standard approach for clinical genetics. The patient phenotypic data is summarised in **Figure S2**. **Table 2** shows the 66 candidate “pathogenic” variants identified, with ACMG/AMP criteria and scores ≥ 10. We note that some genes (*CFHR1, CFTR, IFIH1, IL17F, MEFV*, etc.) have unique variants from multiple patients, supporting their effect. **Figure S3** shows the automated evidence interpretation of these variants. We automated downstream interpretation of evidence uniformly in all of our analysis including ACMG/AMP criteria.

**Figure 1.**
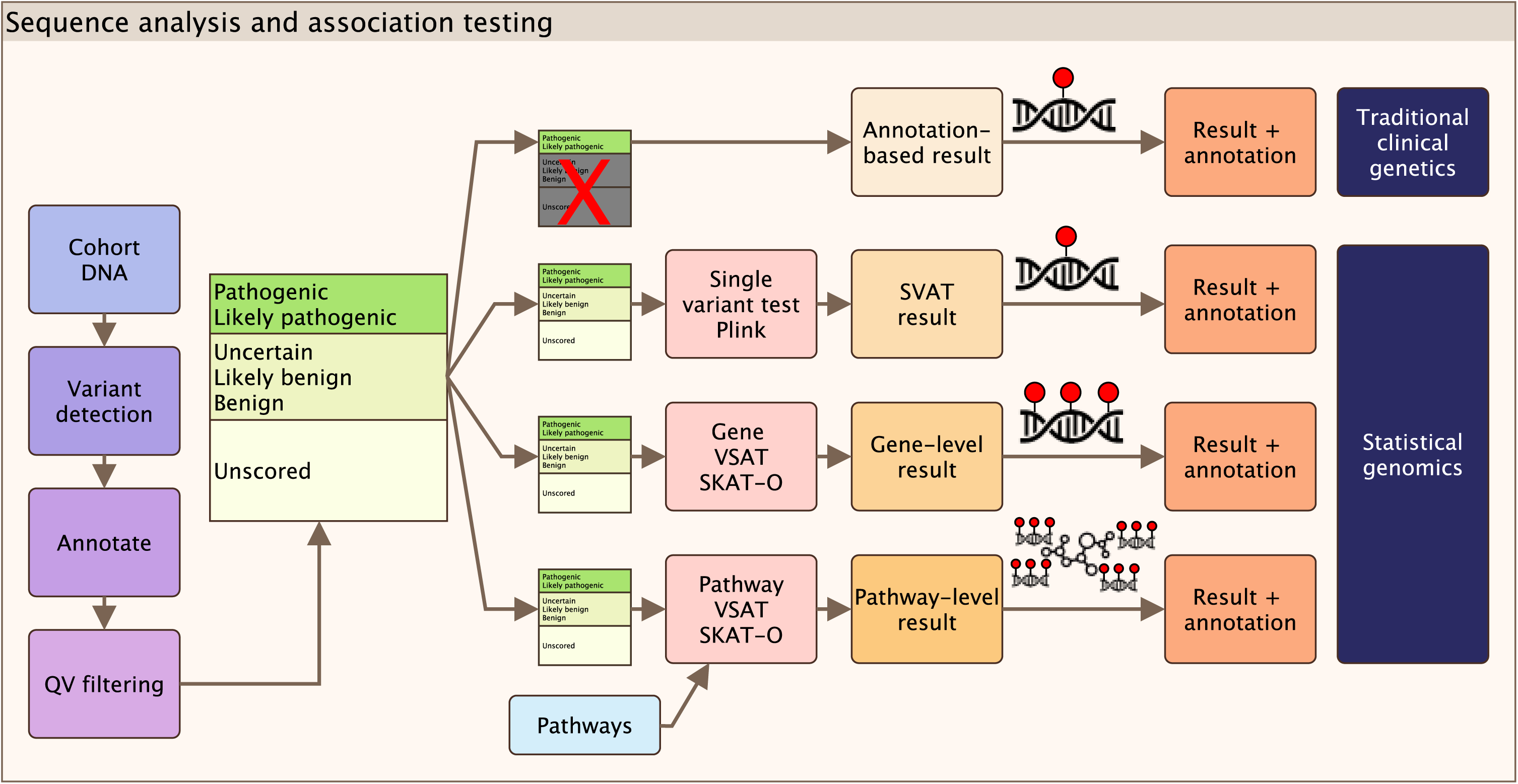
Sequence analysis and association testing overview. Cohort data consisted of exome sequence data, clinical and demographic covariates, and disease status. A single pipeline was used for GATK best practices, annotation with VEP, and quality control to produce a set of qualifying variants (QV). The QV dataset was then used in each of the analysis steps. A clinical genetics report for individual cases was produced based on annotation. Statistical analyses were carried out at the variant, gene, and pathway levels. Results were jointly assessed with Archipelago plot and ACMGuru for variant interpretation. QV, qualifying variant; SVAT, single variant association test; SKAT-O, optimised sequence kernel association test, VSAT, variant set association test.

Single-case analysis integrates variant scoring by necessity using the International Union of Immunological Societies’ (IUIS) Inborn Errors of Immunity (IEI) (527 gene) list (27) (**Table S1, S2**). The IUIS IEI-related genes covered 7786 variants within single-case analysis. An illustration of resulting evidence annotation is shown in **Figure S1** (left) panel 1 (databases in **Table S1** and summarised in **Figure S1** annotation). The filtering protocol was per ACMG/AMP standard (15) (**Tables S3-S8**).

**Figure S3** shows the graphical illustration of known protein functional data for single cases where variants were found and determined as candidate “pathogenic” with scores ≥ 10 (expanded in **Figure S4**). AutoDestructR was used to automate mapping of all variants on protein structures. An example result of individual case summary is shown in **Figure S4 (A-D)**. The full protein pathway for all “pathogenic / likely pathogenic” gene variants was retrieved to provide context as a shared determinant for sepsis within the cohort **in Figure S4 (D)**. The clinical features of the cohort, summarised in **Table 1,** showed no significant association with individual candidate causal variants, as shown in **Figure S5**.

A subset of 176 patients was previously analysed in Borghesi *et al.* (2020) which investigated rare variants in primary immunodeficiency genes in children with sepsis and 11 patients with community-acquired blood-culture positive *P. aeruginosa* sepsis in Asgari *et al*. (2016) (18)(19). Our updated single-case analysis adheres to stricter standardized criteria (15), including scoring methods that were published after the initial study (28). We also incorporated more extensive evidence sources (10), leading to a more selective identification of pathogenic variants. During processing with ACMGuru, we confirmed the presence of the three main variants (*WAS* ENSP00000365891.4:p.Glu131Lys, *CFH* ENSP00000356399.4:p.Pro503Ala, *CYBB* ENSP00000367851.4:p.Gly364Arg) reported in Borghesi’s study. An additional set of 41 variants of unknown significance (VUS) were listed in their supplemental data. However, no additional evidence was found to support these VUS to pass our enhanced evidence thresholds necessary for inclusion as main results. We include an extensive supplemental validation report on the revaluation and updated classification for this subgroup. Although not ranked highly enough for main text inclusion, these variants are still considered as candidate causal for the individual patients, reaffirming the earlier report and remaining relevant for the individual cases. This approach ensures that only variants with strong supporting evidence are highlighted in **Table 1**, reflecting advancements in variant scoring and interpretation.

### 3.3 Single variant analysis

In single rare variant association testing (**Figure 2 A-B**) we did not detect any significantly enriched single rare variant by 1df chi-square allelic test. **Figure 2 (A-B)** shows a Manhattan plot and QQ-plot including the significance threshold after Bonferroni correction for multiple testing, −*log*10(3.97*e* − 07) = 6.40, based on 125,975 single variant tests. Population stratification was controlled by principal components analysis (PCA) based on the variance-standardised relationship matrix, as shown in **Figure S6**.

**Figure 2.**
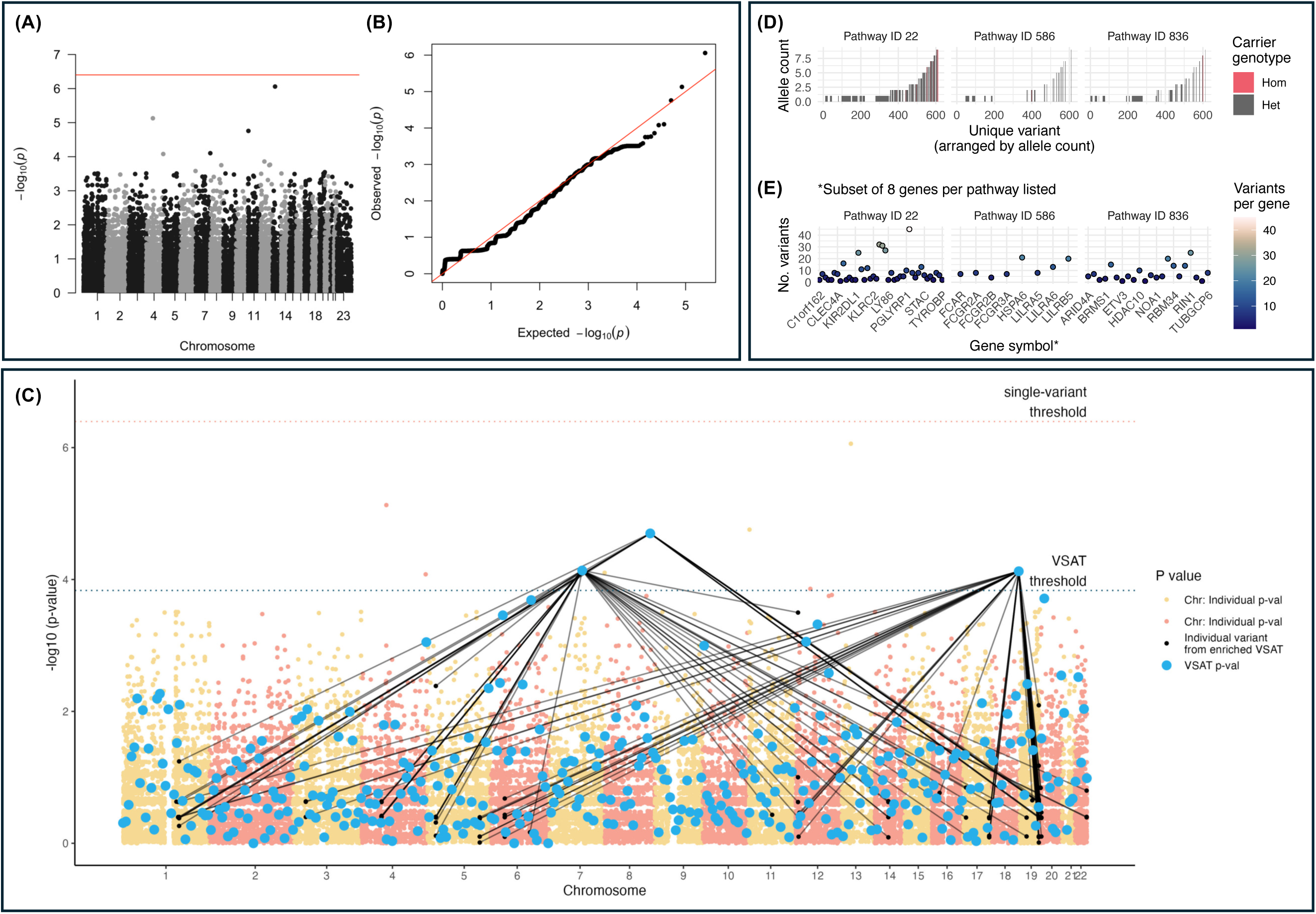
Single variant test and variant set association test (VSAT) with Archipelago plot for enriched protein pathways. Case-control analysis showing (A) Manhattan plot and (B) QQ-plot for single-variant case/control analysis. (C) Archipelago plot showing p-values for case/control association testing in (i) protein pathway VSAT (blue) and (ii) individual variants (yellow and orange points). Each significantly enriched VSAT p-value is mapped to its constituent individual variants’ p-value. The significant threshold based on Bonferroni correction for the number of VSAT is shown by the lower band (blue) and for individual variant tests is shown by the upper band (orange). The three significantly enriched variant set IDs (protein pathways) were pathway 22 and 586 (both later identified as involved in immune response to infection) and pathway 836 (later identified as involved in nucleosomal DNA binding). Raw VSAT p-values are shown in **Figure S8.** VSAT, variant set association test. (D) Distribution of variant carriage within the enriched protein pathway. Homozygous variants are indicated by the red color. (E) The number of unique QV variants in the tested pathway are shown per gene. QV, qualifying variant; 1, heterozygous; 2, homozygous.

### 3.4 Protein pathway clustering

To avoid bias that occurs by fitting pathways during/after statistical analysis, we first constructed protein pathways using external protein-protein interaction (PPI) databases before VSAT analysis (11) using ProteoMCLustR. We used the gene set of pathways for all genome-wide protein-coding genes with known PPI (**Figure S7**). The initial network included 19,566 proteins. **Figure S7 (A-B)** illustrates the resulting pathway sizes ranging from 5 to 50 proteins. **Figure S7 (C)** illustrates the algorithm implementation. The resulting *a priori* constructed network was then used for our pathway-based analyses.

### 3.5 Gene-level and protein pathway-level analysis (VSAT)

Gene-level and pathway-level VSAT was run with variant collapse (13), using SKAT-O with resampling (20),(30) as a case/control study (29),(20),(21). The first ten principal components (PCs) were included as covariates to account for potential population structure (**Figure S7**). We found no significantly enriched genes using VSAT gene-level testing. In pathway-level testing we identified three significantly enriched protein pathways -variant set pathway IDs 22, 586, and 836.

We independently designed and validated the Archipelago R package as a method to summarise and illustrate VSAT results in their genomic context, similar to how the Manhattan plot is used for GWAS. **Figure 2 (C)** shows the result of VSAT in protein pathways. Both single-variant (as per **Figure 2 A**) and VSAT (pathways) results are shown. Each single-variant was mapped to its protein pathway if it had a significant association in VSAT analysis. The raw VSAT p-values, have no natural x-axis position and must otherwise be ranked based on association strength, are reported in **Figure S8.**

The individual variants that contributed to each enriched pathway are shown by black lines in **Figure 2 (C).** Most variants were heterozygous singletons **(D)** thus illustrating why rare variants, by default, are not detected in isolation by GWAS before VSAT. The potential contributions of low and high frequency variants are discussed in supplemental materials. Most genes contributed less than ten unique variants indicating relatively consistent contributions across the pathways **(E)**.

### 3.6 Enriched pathway genes

The enriched pathways consisted of the following genes: pathway ID 22 had *C1orf162, CCDC61, CD300C, CD300E, CD300LB, CD300LF, CLEC4A, CLEC5A, CSF1R, HCST, HMGXB3, IGSF6, KIR2DL1, KIR2DL4, KIR3DL1, KIR3DL2, KIR3DL3, KLRC1, KLRC2, KLRD1, LILRA1, LILRB1, LILRB4, LY86, MNDA, MS4A14, MS4A6A, MS4A7, PDGFRB, PGLYRP1, SIGLEC1, SIGLEC14, SIGLEC7, SIRPB1, SIRPG, STAC, TREM1, TREM2, TREML1, TREML2, TREML4, TYROBP*; pathway ID 586 *had FCAR, FCGR2A, FCGR2B, FCGR3A, HSPA6, LILRA5, LILRA6, LILRB5*; and pathway ID 836 had *ARID4A, ARID4B, B4GAT1, BRMS1, CHD3, CHD4, ETV3, GATAD2A, GATAD2B, HDAC10, HDAC2, MAPK12, NOA1, NOP2, PRDM9, RBM34, RECQL5, REST, RIN1, RNF227, SAP30BP, TUBGCP6*. **Table S9** shows the main annotations for genetic features. We identified a total of 1,113 variant observations (1,101 heterozygous, 12 homozygous) in 418 of 534 patients, of which 601 were unique variants (596 heterozygous, 5 homozygous), 400 of which were singletons. The inheritance pattern of known IUIS IEI were checked with only 7 variants present (autosomal recessive) and 594 unknown. This affected only one IUIS IEI gene (i.e. primary immunodeficiency genes), *FCGR3A*, where 2 of 7 heterozygous variants were of high impact (ENSP00000392047.2:p.Gln70Ter, ENST00000443193.6:c.41-2A>G). The functional variant impact consisted of 45 high impact variants (frameshift, stop gained or lost, start lost, splice acceptor or donor), 556 moderate variants (missense, inframe deletion), and 12 homozygous moderate.

### 3.7 Clinical genetics classification

We analysed variants identified in the significantly enriched protein pathways for evidence of pathogenicity, as detailed in **Table S3-S5** and illustrated in **Figure 3** and **S7**. Figure 3 **(A)** reports the results of ACMG/AMP criteria in evaluating the evidence supporting the pathogenicity of these variants. We prioritised variants supported by existing evidence **(A)**. For in silico predictions, which represent the sole prediction-based criterion among the 25 ACMG/AMP standards, a stringent assessment was conducted (**Figure 3 B**). A variant was only considered to meet the in silico criterion (PP3) if it was supported by at least eight prediction engines. This rigorous selection process ensures that only the most likely pathogenic variants, representing the top 1% of the filtered set, are classified under PP3, as shown in **Table S4**. The ACMG/AMP scoring rules facilitate the integration of pathogenic and benign evidence as documented in**Tables S6** and **S7**. This methodological framework culminates in a definitive ACMG/AMP classification, summarising the combined pathogenic and benign scores, with the final classifications results, as presented in **Table S3**.

**Figure 3.**
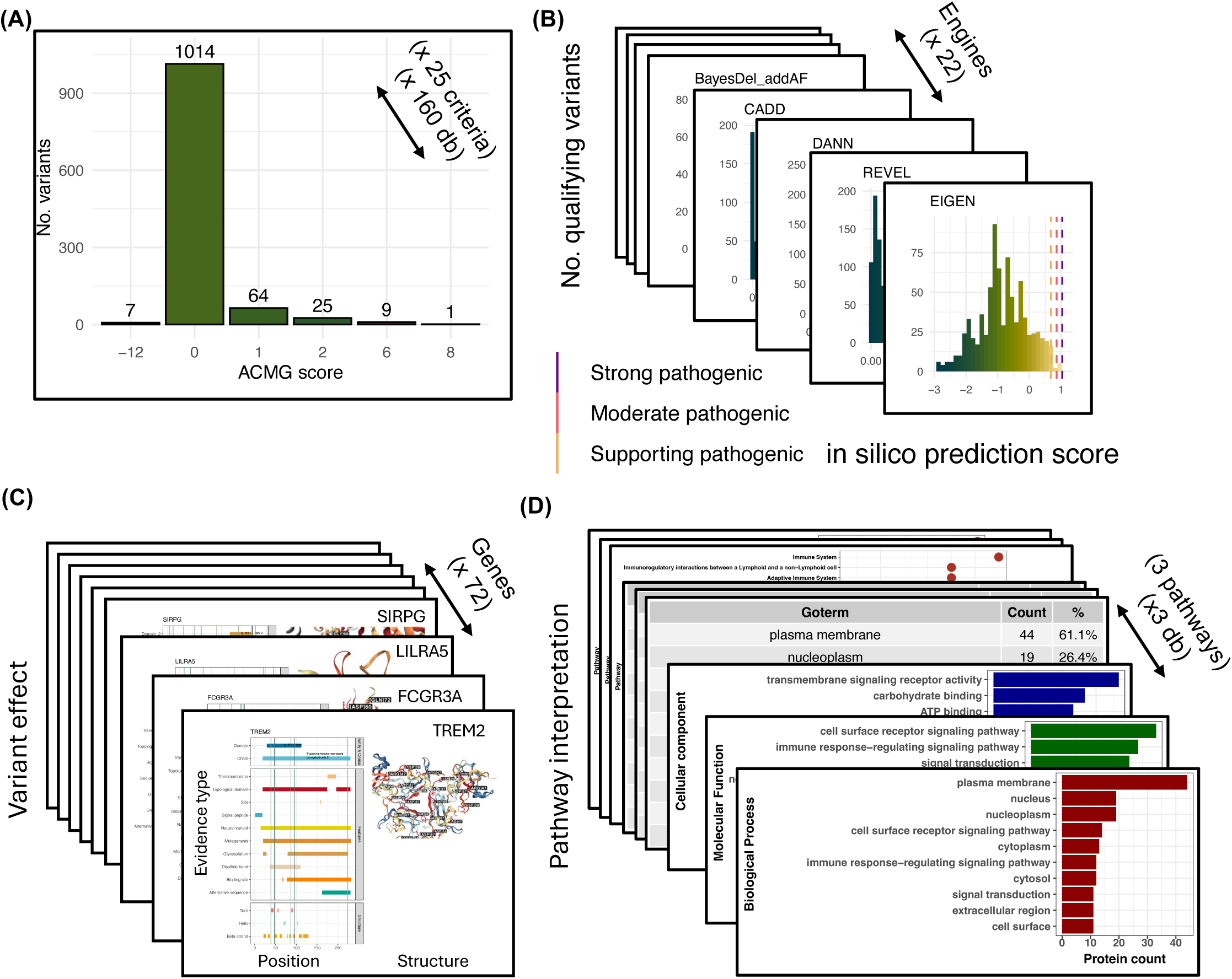
ACMGuru interpretation of pathogenicity in sepsis-associated protein pathways. ACMGuru gene illustrate visualised the output for ACMGuru post PPI containing the enriched pathways 22, 586, 836. (A) The different criteria for evidence interpretation are summarised here to show only the final determination of total score by ACMGuru. (B) Distribution of variants for each of the prediction engines with reference sources for strong, moderate, and supporting evidence. (C) Gene structure evidence plots and protein structures. Variants are illustrated by vertical bars, with density indicating ACMG scoring. (D) Combined KEGG, Reactome, and Gene ontology interpretation of the genes reported in the enriched protein pathways with ACMGuru uniprotR. The fully expanded version of this image is included in supplemental figures.

### 3.8 Predicted impact on protein

The scoring mechanism shown in Figure 3 A-B prioritised candidate causal variants. Structural and functional information supporting variant effect was only then used for interpretation. Figure 3 **(C)** illustrates the significantly enriched pathway-level VSAT for variants that have some known functional effect. Variants are marked by vertical bars at their amino acid positions while the known protein features are illustrated by horizontal bars. **Figure S9** shows the expanded dataset for genes with known effects. AutoDestructR was used as a wrapper for several steps in protein structure visualisation, including NGLVieweR (31), for plotting and annotating PDB structures. Figure 3 **(C)** shows output where we confirmed labelling, measure distances, and other features for interpretation to validate the result evidence from the other sources upstream. **Figure S10** shows an example gene to highlight the interpretation process at the single-sample level. Thus, variants that overlapped with known functional domains were interpretable both automatically and manually, while those without known data were interpreted only for their statistical enrichment from VSAT.

### 3.9 Clinical features and variant count

In our global review of clinical features, we found no significant differences in the characteristics among the case group (548 of 940) who carried variants associated with VSAT enrichment (**Figure S11**). This consisted of 419 carriers of 548 total. The variant carrier breakdown per VSAT pathway was 311 versus 237, 130 versus 418, and 192 versus 356 in pathways 22, 586, and 836, respectively. We measured the overlaps among the variant carriers and found 235 individuals belonged to one pathway, 151 carried variants in two pathways, and 32 individuals had variants in all three pathways. Singleton variants are more likely to have a strong effect in single individuals, and the majority of shared benign variants (which are down-weighted in SKAT-O) contribute little effect. Thus, each individual within the enriched VSAT pathway group is unique while having some shared phenotype specific to their single variant, as represented in **Table S9** and **supplemental report (**automated clinical cohort summary).

### 3.10 Pathway and protein features

The significantly enriched pathway found during pathway-level VSAT was of unknown function. ACMGuru gene-illustrate module to running uniprotR, from Soudy *et al.* (32) resulted in pathway classification. Figure 3 **(D)** (and **Figure S9**) illustrate the interpretation process which showed a strong explanation for two of the protein sets in our enriched VSAT (pathways 22 and 586) as responsible for an immune response to infection, involving antigen presentation and signalling receptors. The main sources, Gene Ontology (33), KEGG (34), and Reactome (35) provided consensus agreement that the identified genes were central to immune cell regulation, antigen processing, and cellular signalling. Pathway 836, which was associated with the regulation of transcription, offered limited interpretative value in relation to sepsis compared to the other pathways.

Figure 4 **(A)** comprises four panels. Panels 1 -3 display the PPI composition for each sepsis-enriched pathway (22, 586, and 836) following clustering and independent enrichment testing. Panel 4 presents the complete STRINGdb PPI network for these enriched pathways - a view that is only possible after the clustering and independent testing have been performed. Notably, panel 4 reveals that pathways 22 and 586 share a largely interconnected network, reinforcing their biological linkage and supporting their independent enrichment in sepsis cases. The results of whole-genome PPI clustering and enrichment analysis by VSAT was illustrated to show the composition of the individual pathways. Figure 4 **(B)** shows the render of immunological responses which are explained by the dysfunction within protein pathways 22 and 586. Approximately half of the genes within the pathways are shown in this render.

**Figure 4.**
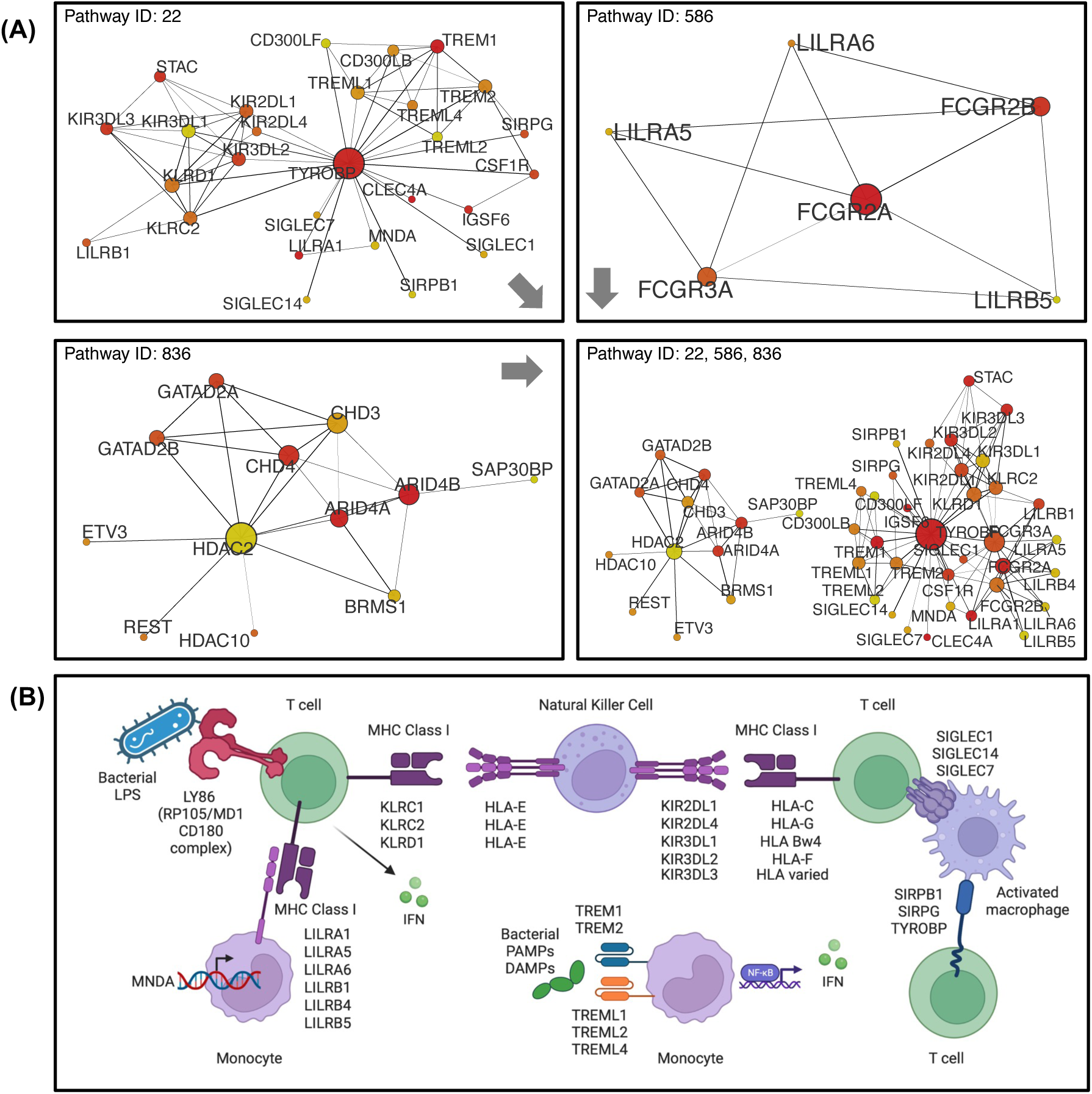
Structural visualization of protein pathways and variant impact in sepsis. (A) Illustration of proteins interactions within their functional pathway. UntangleR used the output of VSAT analysis and merged with data from STRINGdb for 9606 (human) v11.5. Nodes represent proteins and edges indicate known/reported PPI. Node size reflects the degree of connectivity, with larger nodes signifying more interactions and an ostensibly central role in their pathways. The color gradient from yellow to red denotes increasing connectivity, assisting priority to key regulatory proteins. This visualisation elucidates the regulatory hierarchies and interplay among proteins according to existing evidence. (B) Render of protein pathway (*22, 586*) *functionally impacted for patients presenting with sepsis.* .

## 4 Discussion

Our study identified three novel sepsis-associated pathways, encompassing 77 genes, alongside 66 single-case variants of inborn errors of immunity. In keeping with an unbiased, data-driven approach, we have deliberately refrained from focusing on any single causal gene, instead allowing the data to guide our insights. By integrating validation from clinical genetics, gene function, protein structure analyses, protein interaction networks, and population data, our findings deepen our understanding of sepsis’ genetic architecture and provide a robust framework for future genomic investigations of sepsis and other complex diseases.

Understanding the genetic factors that contribute to immune dysregulation in sepsis is a crucial step towards developing precision approaches which can improve outcomes in affected children. The analytical advancements that we developed for this study enhanced our ability to discern genetic contributors to paediatric sepsis. The challenges in such diseases include the potential for both rare and shared variants to cause disease, the application of statistical methods that are appropriate and not biased by targeting subjective gene-sets, and the automated interpretation of both traditional single-case analysis and statistical cohort analysis such that quantification of evidence about biological function is reproducibly enforced.

We employed a multi-level genomic profiling approach that included single-case, variant, gene, and pathway-level analyses. Key to our methodological innovations was the development of ProteoMCLustR, a robust tool for protein pathway clustering. A simple wrapper for SKAT-O allowed us to perform a consistent VSAT analysis on all variant, gene, and pathway levels for high-throughput statistical analysis. A critical advancement was the complete automation of variant interpretation and classification. ACMGuru was thus integral for removing any selective bias. It allowed for consistency during single-case analysis and VSAT analysis. After a blind VSAT - independent of gene or pathway function - the statistically enriched dataset could be automatically interpreted without manual curation, therefore improving the reliability and reproducibility since all supporting evidence is publicly available.

The integration of these technologies facilitates the genetic analysis of complex diseases like sepsis. However, our approach had several limitations. By performing whole exome sequencing (WES), we targeted protein-coding regions, which compared to whole genome sequencing (WGS) offered cost benefits and streamlined analysis but limited our insights into the impact of regulatory variants. In this study, we also did not conduct functional validation on newly identified variants; our computational biology approach relies on existing sources of evidence. We were also limited by the cohort size and by the heterogeneous nature of sepsis - which is likely due to a combination of multiple genetic and environmental factors.

This setup enabled us to identify three enriched pathways. Two of the pathways were implicated in paediatric sepsis, comprising immunoregulatory gene families such as Killer Cell Immunoglobulin-like Receptors (KIR), Killer Cell Lectin-like Receptors (KLR), Leukocyte Immunoglobulin-Like (LIL) receptors, Triggering Receptors Expressed on Myeloid cells (TREM), and Fc Gamma Receptors (FCGR). These gene families encode proteins essential for immune surveillance, cellular signalling, and response modulation during infection.

Considering the protein pathway enrichment overall, these variants are not detected in single-case analyses due to the lack of existing evidence of their pathogenicity. However, when analysed within a combined cohort, each individual variant contributes to a previously defined pathway, potentially resulting in a shared phenotype. For the pathway analysis only one IUIS IEI gene was affected, FCGR3A, where 2 of 7 heterozygous variants were of high impact. In the pathway-based analysis 419 of 548 individuals were involved for the enriched pathways, although most variants will contribute little effect. In the single-case analysis 63 patients and 66 variants were identified. There was an overlap of 54 patients resulting from both analysis. We found no particular clinical feature to be significantly associated with individual candidate causal variants (**Figure S5)**, indicating that decision rules based on clinical features such as age or severity to guide genomic investigation in children with sepsis are unlikely to be meaningful. However, this does not rule out the potential of more complex multivariate predictors in a clinical setting.

Our analysis focused on a rigorous statistical approach. Any subsequent functional validation of these newly identified protein pathway-based associations will require focusing on a selection of top candidates. We highlight a few candidate pathogenic variants selected from the top evidence scores. Notably, in pathways associated with sepsis, truncating variants in *KIR2DL4* and *KIR3DL3* are thought to disrupt signal transduction, and a single-nucleotide variant in *KIR2DL4* which reportedly disrupts the signal peptide (36), all of which are predicted to impact the receptor’s ability to modulate immune tolerance and possibly contributing to sepsis through an imbalance in NK, CD8+ T cells, and B cell functions (37), (38). Signalling occurs via *KIR2DL4* as the receptor for HLA-G complex which is linked to septic shock in critically ill patients (39), further illustrating the vital role of these signalling processes in immune responses.

Moreover, loss-of-function (LoF) variants that we detected in *KIR3DL3* and *KIR3DL1* may impair NK cell-mediated cytotoxicity and immune surveillance, allowing pathogen proliferation and increasing sepsis risk. This potential mechanism is supported by phenotypic shifts towards inhibitory receptors and reductions in NK cell cytotoxicity and activation markers CD69 and CD107a during sepsis (40), suggesting that carriers of these variants might experience similar immune dysfunctions (41). The tolerance of heterozyous LoF is difficult to quantify for sepsis susceptibility and thus warrants further investigation to determine its impact (42).

Additionally, we found LoF variants in *LILRA1* affecting glycosylation and Ig-like regions, potentially disrupting immune signalling (43). Loss-of-function variants detected in *LILRB1*, which regulates phagocytosis, could similarly influence sepsis severity by impairing immune responses. While this mechanism is currently a promising target for cancer immunotherapy, LoF during pathogenic infection could worsen the potential for sepsis (44).

Our analysis also identified variants in *SIGLEC1*, including seven LoF and two missense variants, that could hinder pathogen clearance and exacerbate sepsis severity, paralleling findings in other SIGLEC family genes which are already recognised as a novel therapeutic approach in sepsis (45). A similar profile was present for *SIGLEC7* and *SIGLEC14*. Notably, *SIGLEC5* has been proposed as a sepsis biomarker, though it was not found in this cohort, has functionally similarity to the members detected here (46). Lastly, we detected LoF variants in *SIRPG* which was one of eight genes identified that most accurately predicted early onset sepsis (EOS) and differential gene expression in EOS infants suggests potential targets for precision medicine (47).

This brief sampling of top candidates underscores the importance of exploring these molecular pathways and variants for their roles in sepsis pathophysiology and their potential as therapeutic targets. The exact mechanistic links between our reported variants and sepsis susceptibility, disease progression, and outcome must be tackled in multiple steps: (1) conducting individual mechanistic studies to test function; and (2) generating and replicating new disease association datasets, which can act as early warning signals in future. In the first step, our results demonstrate the prioritised variants of interest both at the cohort level and single-case level for follow-up functional assays and for testing in biomarker development. In the second step, replication studies in new cohorts with broader demographics could further reinforce the association or causality of these genes and pathways with sepsis susceptibility.. By design, our protocol is agnostic to the disease phenotype, enabling straightforward application to new cohort studies of rare diseases.

Given the systemic and intricate nature of sepsis, our study suggests that the impact of individual damaging gene variants, each manifesting via unique mechanisms, coalesces into a shared phenotype. This commonality is rooted in the affected protein pathway that is consistent across all patients, ultimately leading to a dysregulated immune response, a primary hallmark of sepsis. Further research is needed to understand these complex interactions and their implications for sepsis pathogenesis and treatment. These findings may shed light on the molecular mechanisms underlying this condition and pave the way for the development of more effective diagnostics and therapeutic strategies for paediatric sepsis.

## 5 Conclusion

We developed and applied a novel bioinformatic pipeline to explore the genetic basis of sepsis susceptibility in children, using a large cohort with blood culture-confirmed sepsis. We identified three critical protein pathways of 77 genes encoding proteins involved in antigen presentation, cellular signalling, and immune cell interactions. We additionally identified 66 variants in individual cases. Our approach not only elucidates the complex genetic underpinnings of sepsis but also sets the stage for future investigations that may lead to more targeted and effective treatments.

## Data Availability

All summary statistic data are present within the work. Raw genomic data is controlled under restricted access according to ethical guidelines. All code is published.

## 7 Acknowledgements

We thank the participating patients and their families; and the study coordinators at each study site. We thank Lars Malmström for work his as an independent reviewer.

## 8 Ethics statement

The study was approved by the respective ethics committees of all participating centers (Cantonal Ethics Committee Bern, approval number KEK-029/11) and the study was conducted in accordance with the Declaration of Helsinki.

## 9 Authorship Contributions

Dylan Lawless performed analysis, designed methods, and wrote the manuscript. Philipp Agyeman collected and managed data, performed analysis, and wrote the manuscript.

Pauline Rogg, Manon Bouzereau, Robin Fallegger, Zaira Seferbekova, Valeriia Timonina, Konstantin Pipadin, Ali Saadat, Zhi Ming Xu, Christian W. Thorball, Flavia Hodel, and Johannes Trückk designed methods. Jacques Fellay and Luregn J Schlapbach designed the study, supervised work, and wrote the manuscript.

## 12 Abbreviations

ACAT: aggregated Cauchy association test
ACMG/AMP: American College of Medical Genetics and Association for Molecular Pathology
ClinGen: Clinical Genome Resource
GATK: Genome Analysis Toolkit
GLM: generalised linear model
gnomAD: genome aggregation database
GRM: genetic relationship matrix
GWAS: genome wide association study
HPC: high performance computing
IEI: inborn error of immunity
IEM: inherited errors of metabolism
IUIS: International Union of Immunological Societies
LD: linkage disequilibrium
LoF: loss of function
MCL: Markov cluster algorithm
MDS: multidimensional scaling
MHC: major histocompatibility complex
NK: natural killer
PCA: principal component analysis
PM3: pathogenicity moderate 3
PPI: protein pathway interaction
PS1: pathogenicity strong 1
PVS1: pathogenicity very strong 1
QC: quality control
QV: qualifying variant.
SCITAS: Scientific IT & Application Support
SKAT: sequence kernel association test
SKAT-O: SKAT optimal unified
SNV: single nucleotide variant
STAAR: variant Set Test for Association using Annotation infoRmation
SVAT: single variant association test
VEP: Variant Effect Predictor
VSAT: variant set association test.
VUS: variants of unknown significance
WES: whole exome sequencing
WGS: whole genome sequencing.

## 13 Tables

**Table 1 Demographic and clinical characteristics children with blood culture-proven bacterial sepsis**

Categorical variables are presented as frequencies (%) and continuous variables as median, min-max, SD. Column percentages are presented for total cohort cases.

**Table 2 Single-case variants classified as pathogenic or likely pathogenic.**

This table contains the key clinical data measurements for each patient including the major genetic feature annotations for all variants that were classified as pathogenic or likely pathogenic. AD: autosomal dominant; AR: autosomal recessive; XL: X-linked. Death or extended PICU is composite endpoint indicating whether the child either died within 30 days after blood culture sampling or had a PICU stay of ≥3 days. PODIUM score is a composite metric summarising sepsis severity where lower scores indicate milder clinical status, whereas higher scores reflect greater severity and risk of adverse outcomes.

## 16 Links

Full study code repository: https://github.com/DylanLawless/spss_exome_vsat

ACMGuru repository: https://github.com/DylanLawless/ACMGuru

ProteoMCLustR repository: https://github.com/DylanLawless/ProteoMCLustR

Archipelago repository: https://github.com/DylanLawless/archipelago

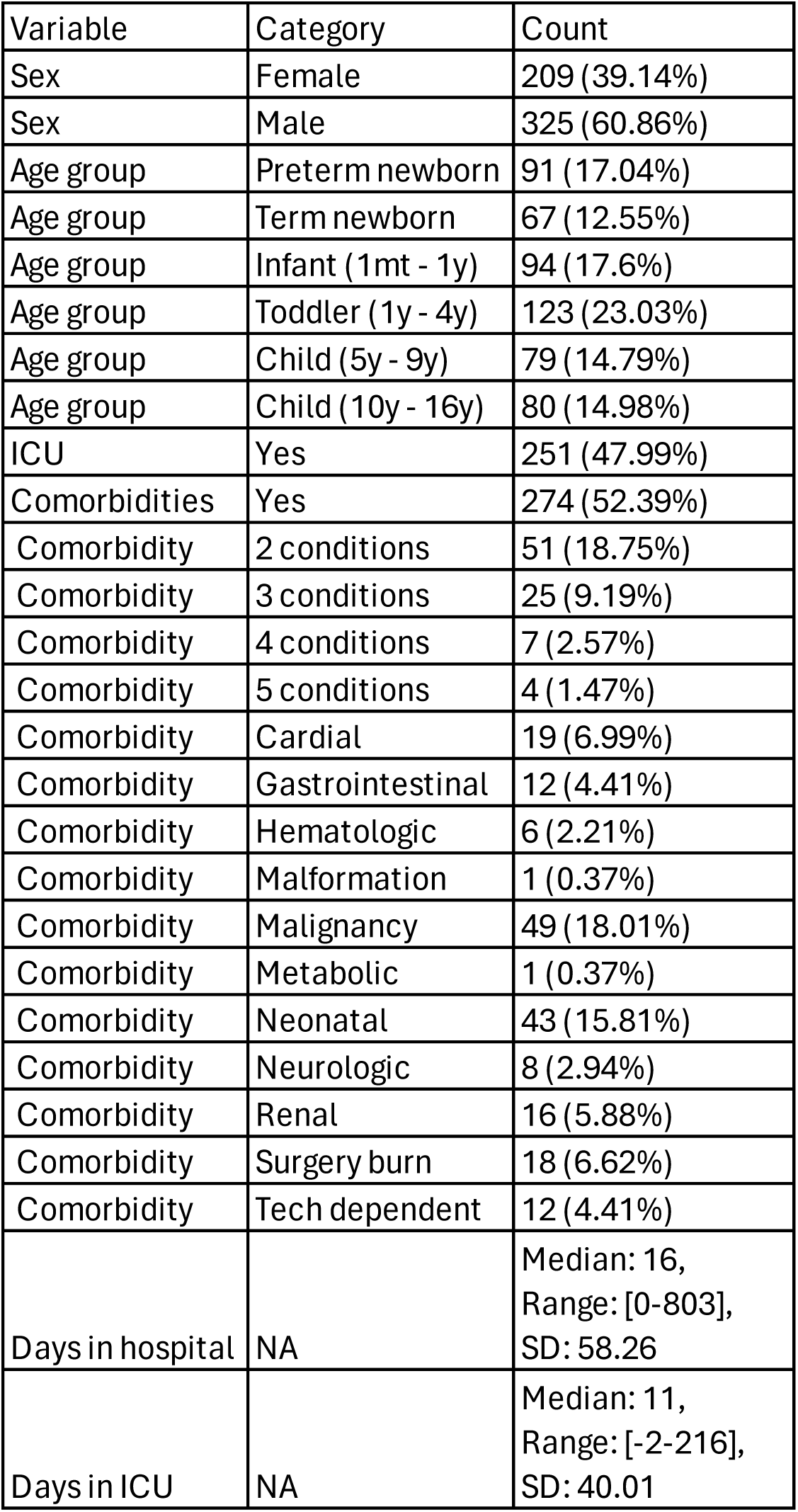

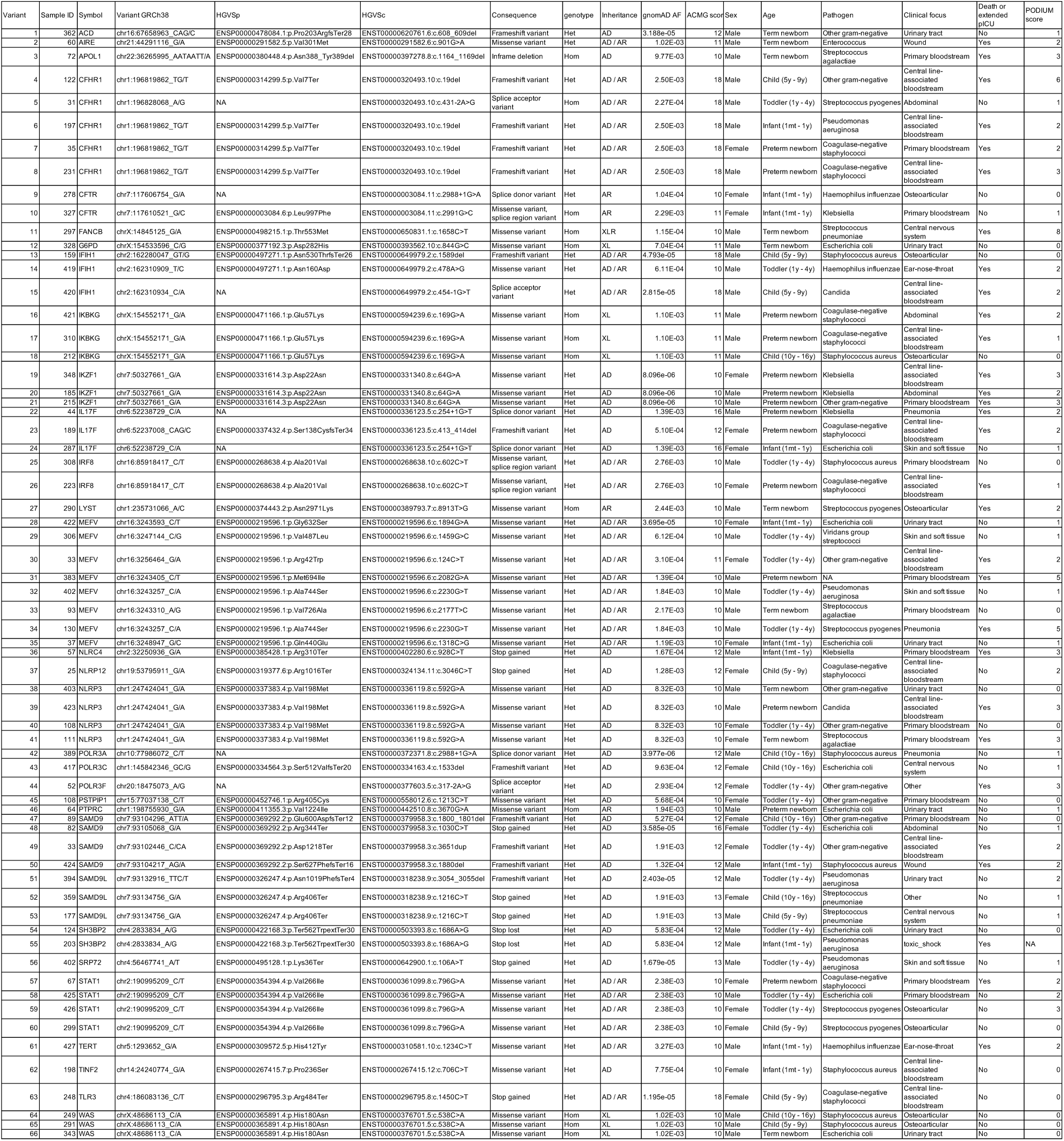

## 1 Online methods

### 1.1 Sample collection and exome sequencing

Children under 17 years with community-acquired or hospital-acquired blood-culture positive sepsis were eligible. This consisted of 534 cases, DNA from 14 parents, and 392 controls. Details of the cohort have been previously reported (1)(2). A subset of 176 patients was previously analysed in Borghesi *et al.* (2020) (3) which investigated rare variants in primary immunodeficiency genes in children with sepsis and 11 patients with community-acquired blood-culture positive *P. aeruginosa* sepsis in Asgari *et al*. (2016) (4). The in-house controls consisted of 392 subjects from the Swiss population. The control cohort was 76.79% adult male (301 of 392), therefore our analysis included statistical controlled for age and sex and was restricted to germline genomic variation. This study was carried out in accordance with the recommendations of the ethics committees of participating centers. Parents/guardians of all included patients gave written informed consent in accordance with the Declaration of Helsinki.

### 1.2 Exome sequence preparation

Blood was drawn from patients who had agreed to participate in the study following a positive blood culture notification to medical staff. Blood was collected for serum and DNA analysis (1-2 ml each, using EDTA tubes), with the processing details as previously published (4). DNA was extracted from the collected blood following the protocol provided by the manufacturer (QIAamp DNA blood kits, Qiagen, Crawley, UK). Whole Exome Sequencing (WES) was then carried out, starting with cluster generation using Illumina TruSeq PE / SBS Cluster Kit v5 reagents and Covaris E220 Focused ultrasonicator. The libraries were sequenced to produce 100 / 150 bp paired-end reads on an Illumina HiSeq 2500/3000 (Illumina, San Diego, CA, USA) achieving a minimum coverage depth of 30X. The sequencing output was processed using Illumina CASAVA v1.8.2 software for raw image analysis, base calling, and FASTQ file generation.

### 1.3 Computing environment

All analysis, downstream of sequencing, was performed on the EPFL community Scientific IT & Application Support (SCITAS) high performance computing (HPC) platform using the Intel Ice Lake based cluster. This was composed of 419 compute nodes, each with 2 Intel(R) Xeon(R) Platinum 8360Y processors running at 2.4 GHz, with 36 cores each (72 cores per machine), 3 TB of SSD disk, which have a minimum of 512 GB of RAM per node. The maximum requirements for any single analysis step included time: 12 hours; 24 CPUs per node; 80 GB per CPU; an array of approx. 1000 jobs; for approx. 1000 samples. The resulting runtimes for 1000 sample WES analysis were approx. *<* 8 hours for the GATK pipeline, *<* 10 hours for protein pathway clustering (parallel to GATK), and *<* 2 hours for statistical analysis.

### 1.4 Sequence analysis

Exome sequence data in fastq format were trimmed and checked for quality control using fastp v0.23.4with its default and metrics options, respectively (5) (github.com/OpenGene/fastp). For each sample, reads were aligned to the GRCh38 reference genome (GCA 000001405.15 GRCh38 no alt analysis set.fa.gz sourced from: ftp://ftp.ncbi.nlm.nih.gov/genomes/all/GCA/000/001/405/GCA_000001405.15_GRCh38/ seqs_for_alignment_pipelines.ucsc_ids/GCA_000001405.15_GRCh38_no_alt_analysis_set. fna.gz) using BWA-MEM v0.7.17 (6) and converted to BAM format using SAMtools v1.14 (7). GATK v4.2.2.0 best-practices for short germline SNV/Indel were used for marking duplicate reads (MarkDuplicatesSpark), variant recalibration (BaseRecalibrator and ApplyBQSR), outputting genomic VCFs with HaplotypeCaller (HaplotypeCaller) in either/or ERC base pair resolution mode and ERC gVCF mode, and joint genotyping (GenotypeVCFs) (8). We included standard quality control (QC) filters on the joint genotyped dataset. Recalibration was performed with hapmap 3.3.hg38.vcf, 1000G omni2.5.hg38.vcf, 1000G phase1.snps.high confidence.hg38.vcf, Mills and 1000G gold standard.indels.hg38.vcf, dbsnp 146.hg38.vcf, truth sensitivity filter level 99.7 for SNP and 95 for INDEL, refinement with genotype posteriors, flagging low GQ < 20. After exploration for quality, we performed further filtering: QUAL≥ 30, INFO/DP ≥ 20, FORMAT/DP≥ 10, FORMAT/GQ ≥ 20. We performed normalization and decomposition to break multiallelic sites using VT (9). We then began filtering based on cohort frequency for the analysis required, such as max-maf 0.01, max 5 homozygous carriers, max 10 heterozygous carriers, etc. in cases/controls combined. Downstream filtering and prediction of functional consequences was performed by annotating data using VEP and a number of database sources as described in later sections. Filtering used either VEP-filter or ACMGuru R scripts during statistical analysis.

### 1.5 Principal component analysis

GCTA was used for constructing genetic relationship matrix (GRM) and principal component analysis (PCA) (10). Due to the presence of long regions of high linkage disequilibrium (LD) in the human genome, it is useful to exclude these regions when performing certain analyses such as PCA on sequence data (11,12). We used the list of positions for GRCH Build 38, provided by Anderson et al. (13) which can be found at https://genome.sph.umich.edu/wiki/Regions_of_ high_linkage_disequilibrium_(LD). The joint genotyped cohort VCF was converted to PLINK format with PLINK v1.9, then filtered to exclude these regions. The first 10 PCs were used as covariates in also association analysis.

### 1.6 Variant filtering

A detailed illustration of the analysis pipeline is shown in **figure 1**, with the top left panel 1 focused on germline variant discovery, annotation, and filtering. After joint genotyping, variant quality score recalibration, and refinement with GATK we had: 716,768 unique variants; after filtering for quality with BCFtools we had: 664,741; after filtering for cohort level MAF to remove common variants we had: 557,256; after VEP annotation and filtering for variant impact we had: 245,088; after filtering based on gnomAD AF < 0.2 (first pass) we had: 201,874. We then split the data into chromosome-level files for parallel computing using SkatRbrain. For any variant, gene, or pathway where statistical significance was detected, the variant set ID (e.g. protein pathway) was used to extract that subset for downstream interpretation. Therefore, the largest filtering step occurred within in VSAT where we reduced the test set to variants within the current gene/pathway.

### 1.7 Sub-setting and variant collapse

Our data was prepared in cohort batches with all cases and controls. Joint genotyping and downstream analysis with GATK best-practices was applied to the joint cohort. Statistical analysis required preparation from the joint cohort dataset depending the computational resource requirements, as follows.

Single-case analysis consisted of annotating and interpretation of all variants in the joint dataset together and only in the final stages, we reported variant carriers where a variant passed our filtering criteria. Therefore we jointly analysed the fully annotated dataset (GATK to VEP with all default and custom plugins). ACMGuru was applied to retrieve final variant results. Single variant testing was performed from the same joint data, however, plink v1.9 was used to convert from VCF to bed, bim, fam format. The joint data was also used as input for gene-level and pathway-level testing. Variant collapse was applied as per Povysil et al. (14) to produce variants set IDs at the gene-level and pathway-level during each of those tests. This step was performed using SKATRbrain, once for each variant set, to retrieved data from chromosome-split files, with annotation from a minimal set of plugins, to reduce the memory requirements.

Significantly enriched variants, genes, or pathways, were then retrieved. As per single-case analysis, ACMGuru was applied to return the final variant interpretation data for each result. The relevant details from each output were passed to Archipelago for a visual summary (p-value, study type, gene symbol, pathway (variant set ID), chromosome, position, and variant ID). Covariates used in statistical analysis were the same throughout, all derived from the upstream joint dataset.

### 1.8 Single variant testing

Single variant testing was performed on the same joint data, after conversion with Plink v1.9 to bed, bim, fam format. MAF filtering was set to 0.01 prior to conversion. The genotyping rate was set to 0.05 to remove variants with missing rates exceeding this value. The Hardy-Weinberg equilibrium (HWE) value was set to 1e-6 to remove all variants which have HWE exact test p-value below this threshold. Covariates consisted of PC1-10, sex, age days^2, study site, and ICU stay. We used plink v1.9 to perform single variant association testing using --assoc and other modes.

### 1.9 Protein pathway construction

A protein pathway interaction (PPI) network was generated to determine a “best fit”, balancing PPI evidence confidence scores to maximise the number of retained genes and pathways, for all proteins genome-wide based on independent reference database before applying it to the cohort dataset. Details and an R package for performing this step, ProteoMCLustR, is described in the extended supplemental methods. Briefly, protein interaction data was sourced from several databases as compiled by STRING database (15). The STRING db is a comprehensive resource for PPI, which integrates both curated data sources - GO, KEGG, Reactome, Biocarta, and BioCyc, and experimental sources - BIND, DIP, GRID, HPRD, IntAct, MINT, and PID. Iterative clustering was run using MCL by ProteoMCLustR until all proteins were assigned to their protein pathway at each of the three recommended PPI evidence confidence scores - 0.4, 0.7, and 0.9 (15,16). Finally, we used the resulting PPI network for the optimal set. A confidence score of 0.7 retained the largest number of proteins assigned to pathways. This provided the variant set ID for each pathway as used during VSAT with SKAT-O to perform case-control analysis, thus analysing every protein pathway, genome-wide. The protocol is summarised in **Figure S5** and detailed steps are further provided in the extended supplemental methods.

### 1.10 VSAT significance testing

SKAT and its optimal unified version SKAT-O are now popular methods for gene-based association tests, accommodating multiple variants within a gene or variant set while accounting for their potentially differing directions and magnitudes of effects. We previously compared a range of association testing methods on synthetic data to assess performance of methods including simple logistic regression, Fisher exact test, logistic regression, CMC, weighted-sum test, SKAT, SKAT-O, etc. Our testing agreed with the recommendations previously reported, finding the SKAT-O methods best suited for this disease study type as demonstrated by Lee et al. (17). SKAT-O is based on the original implementation of SKAT by Wu et al. (18), and demonstrated by Lee et al. (19) and Ionita-Laza et al. (20). Nicolae (21) has reviewed many of the additional similar methods, although none were more appropriate for our usage. We also applied the resampling method for calibrating single and gene-based rare variant association analysis in case–control studies by Lee et al. (22). The p-value calculation uses Davies’ method or a fast moment matching algorithm (23–26). Hence, the *p*-value of *Q*_optimal_ can be quickly obtained analytically with the use of a one-dimensional numerical integration. Small sample sizes (*<* 2000) adjustments are applied accordingly as part of the SKAT R package for our cohort (n = 940). To calibrate the tests and adjust the Type I error rate, the SKAT and SKAT-O methods often employ a resampling procedure. The method by Lee et al. (22) is implemented in the R package SKAT, which we used for SkatRbrain. We expected that single variants would show minimal or no enrichment due to the inclusion of only rare variants. For the same reason, we expected few or no enriched genes or protein-pathways, especially when resampling methods are efficient for controlling type I error Lee et al. (22). We expected any significantly enriched signals to be relevant to susceptibility to infection, although no *a priori* requirements, filters, or selections were set. For any significantly enriched signal, our follow-up consisted of annotation-based interpretation.

### 1.11 ACMGuru: Variant interpretation

#### 1.11.1 ACMGuru variant classification

In our study, we designed and used the ACMGuru R package, for automated interpretation and reporting based on ACMG/AMP guidelines from Richards et al. (27), to classify variants according to standardized and widely recognized rules (**Table S2 – S3**). This allowed us to ensure reproducible and uniform interpretation of variant evidence in all tests. Our annotation process involved using VEP with both default and custom plugins, including dbNSFP, to score and interpret variants. This included generating summary statistics, text descriptions, and various plots, such as variant distributions within genes and pathways, and mapping data onto gene or protein structures from sources like UniProt. We used the default ACMG/AMP classification criteria and followed the recommended methods for updates where necessary, employing stringent upstream filtering and excluding certain ACMG categories like PS2 and PS4 when inappropriate. Variants were classified with specific criteria such as PVS1 for null variants matching disease inheritance, and PS1 for identical amino acid changes to known pathogenic variants. Extensive data sources from more than 160 databases were systematically assessed, including ClinVar, ClinGen, GnomAD. Pathogenic potential was also assessed through multiple computational predictions, including calibrated insilico thresholds from VarSome germline classification, by Saphetor SA, for commonly used metrics like CADD and PolyPhen, applying stringent frequency cutoffs to maintain a conservative approach to pathogenicity assignment. Detailed descriptions are provided in the extended supplemental methods.

#### 1.11.2 ACMGuru gene-illustrate

ACMGuru gene-illustrate uses the UniProt database to enrich our understanding of gene variants. Specifically, we use organism-level data in GFF format, sourced from UniProt (https://www.uniprot. org/uniprot/?query=*&fil=organism%4A%22Homo+sapiens+%28Human%29+%5B9606%5D%22#) Homo sapiens (Human) [9606]), which includes 204,185 entries detailing attributes such as gene and protein names, sequence length, and more. For each variant identified in ACMGuru, gene symbols and UniProt seqid IDs are cross-referenced to ensure analysis focuses on relevant UniProt entries. Variants contributing to significantly enriched variant sets are visually represented by vertical bars at corresponding amino acid positions on the protein models, enhancing the interpretability of functional domains.

The create-plot function within ACMGuru automates the visualisation process, generating unique layout illustrations for each gene or protein, accounting for distinct protein features and experimentally tested variants. This function employs a set of design rules to ensure visual clarity, such as distinct marking of start and end points of protein features, which range from single amino acids to extensive domains. Visualizations are organized with logical groupings on the y-axis to facilitate easy interpretation, while the x-axis consistently denotes the protein amino acid positions.

For broad analyses, including single-case and pathway-level evaluations, the create-and-save-plots function optimises layout dimensions to accommodate the number of protein plots, aiming for a near-square configuration (e.g., 2x2, 3x3) when possible. Adjustments are made when the number of plots does not allow a perfect square layout. Color selections, sourced from the wesanderson R package, are used purely for aesthetic enhancement and do not convey quantitative data.

#### 1.11.3 ACMGuru uniprotR

The outputs from ACMGuru gene-illustrate are automatically passed to ACMGuru uniprotR. This module in ACMGuru directly loads the default UniprotR package from Soudy et al. (28). The default functions of the UniprotR package are used but we include this feature to provide a single workflow by eliminating the manual processing between tools. The uniprotswiss IDs output from the previous step are used, after connecting to the UniProt API, to retrieve data from Gene ontology, KEGG, Reactome, and other sources which are compiled by UniProt. Information about the protein pathway is retrieved for use in interpreting the function of the protein pathway, clustered by ProteoMCLustR and detected as significantly associated with disease using VSAT by SkatRbrain.

#### 1.11.4 ACMGuru get-discussion

To aid in automated report generation, a get-discussion step provides a final CSV or TSV format table to provide context around all of the previous technical descriptions of variants, genes, and pathways by returning the gene symbol list, full gene name, molecular function, and functional description from UniProt.

#### 1.11.5 ACMGuru AutoDestructR

We designed AutoDestructR as an wrapper for several steps in protein structure visualisation, to automatically deconstruct the sets of PDB structures for every gene query and produce an annotated structure image in R. First, it runs ACMGuru to automatically import the results from the analysis chain. Next, biomaRt is called to retrieve data from Ensembl Mart for hsapiens gene ensembl, then using SIFTS database it retrieves pdb chain uniprot data to allow for mapping between uniprotswiss IDs and PDB structure files. Next, since there may be a large volume of partial structures for every gene of interest, we facilitate automation by defining a “best representation” for a given protein. We select representative protein chain records by the selection criteria: (i) maximum chain length, (ii) maximum count of unique PDB entries per protein, (iii) maximum count of unique chain entries per PDB. This retains the structure with greatest coverage and with one or more chains. While the result may not detect the representation with highest resolution or accuracy, in general it results in the best graphical representation. We include an option to output every available structure, should it be required. Lastly, AutoDestructR runs the NGLVieweR package by van der Velden (29) for plotting and annotating PDB structures to output a html format R Shiny plot and png snapshot on transparent background for automated reporting.

### 1.12 Network plotting

We developed untangleR to visualize protein pathway networks using the igraph R package from Csardi and Nepusz (30). Node sizes in the network graphs are scaled according to their connectivity relative to the overall network density, which is calculated as the ratio of the actual to the maximum possible number of edges. Node coloration, termed ’degree colors’, varies from red (most connections) to yellow (fewest connections), reflecting each node’s connectivity degree.

The network visualization employs the ’stress’ layout method based on multidimensional scaling (MDS), positioning nodes to mirror their relational distances within the network. This method effectively highlights the significance of proteins, with highly connected nodes often indicating key roles in cellular functions. Even proteins with fewer, high-confidence interactions can be crucial, particularly in diseases like monogenic rare diseases, where a protein’s selective interactions can significantly influence disease phenotypes.

### 1.13 Single-case variant carrier clinical feature analysis

To see if any clinical features were associated with carriage of variants with evidence of pathogenicity (ACMG score ≥ 4), we adopted two different statistical tests for categorical and continuous variables, respectively: the chi-square test and the Kruskal-Wallis test. We aimed to favour robustness over exact tailoring since we apply this analysis over clinical measurement data which is updated frequently to thus produce standardised procedures which are flexible. In this cohort we had twenty-two categorical and twenty-two continuous variables. The chi-square test can handle more than two categorical variables and test associations across them. The test was performed for each categorical variable against the group, thus allowing us to quantify the degree of association and identify significantly associated categories. It is worth noting that the chi-square test assumes that the categories are mutually exclusive and that the observations are independent. For our continuous variables, we applied the Kruskal-Wallis test, a non-parametric method, to test whether samples originate from the same distribution without making assumptions about the specific distribution type. This makes it particularly useful when dealing with data that are not normally distributed, which was the case in our dataset. It essentially examines whether the median of two or more groups differ significantly. To ensure the validity of our analysis, we excluded observations with missing values from both tests. This complete case analysis was appropriate given that missing values were randomly distributed and constituted a small fraction of our data.

## 1 Supplemental results

### 1.1 Validation of previously reported variants

#### 1.1.1 Summary

In an earlier study by Borghesi et al. (2020), 176 patients were analysed to identify rare genetic variants in genes associated with primary immunodeficiency in children who have sepsis. Our current analysis is greatly expanded in size, uses more stringent criteria based on updated standards (Richards 2016), and scoring methods introduced after the initial study (Tavtigian 2020). We also used additional evidence sources (Liu 2020) which allowed for more selective identification of pathogenic variants.

Using ACMGuru, we re-confirmed the presence of three critical variants: *WAS* p.Glu131Lys, *CYBB* p.Gly364Arg, and *CFH* p.Pro503Ala, initially reported by Borghesi et al. Furthermore, we reviewed additional variants of unknown significance (VUS) which were screened, and those reported in their supplemental data. Unfortunately, no new evidence was sufficient to classify these VUS as pathogenic under our stricter evidence thresholds.

This analysis prompted us to consider remaining uncertainties and the potential implications for future research involving similar cohorts.

#### 1.1.2 Extended

In Borghesi et al.’s earlier study, the variant filtering strategy incorporated allele frequency, CADD scores, population frequency, and a selection of known primary immunodeficiency (PID) genes. Our updated approach used similar foundational elements but enhances the scoring and interpretation process. We now apply a large control cohort, expanded cases, 25 specific scoring rules developed by Richards in 2015 and updated with scoring mechanisms by Tavtigian in 2020, and we use databases that collectively draw from approximately 160 sources (Liu 2011, Liu 2020).

Despite these enhancements, the variants of unknown significance (VUS) identified in the prior study remain in our detection dataset. However, they received low scores on subsequent evidence interpretation because they lack supporting evidence of pathogenicity, despite being associated with PID genes. Figure S3 (A), plot 3, incidentally visually represents this by showing the number of American College of Medical Genetics and Genomics (ACMG) criteria assigned to each variant, illustrating how these new scoring rules impact the classification of variants.

In the Borghesi et al. (2020) study, Table 3 highlighted three specific variants (*WAS, CYBB, CFH)* and Supplemental Table 4 detailed 41 variants of unknown significance (VUS) across 25 genes. In our study, variant detection was followed by qualifying variant (QV) filtering for initial variant identification, resulting in 5780 QV. Using ACMGuru, these candidates were further annotated and split on conditions such as inheritance pattern for (known disease genes) to provide an unfiltered set of 6848 variants. We specifically screened the variants identified in the previous study to assess their presence and significance in our expanded dataset. This approach allowed us to reassess previously reported variants with updated analytical tools and a broader genetic context. We re-identified the main three variants reported previously, now with GRCh38 coordinates and classification scores as shown in Table 10.

The three “known pathogenic” variants were previously reported as *WAS* p.Glu131Lys, *CYBB* p.Gly364Arg, *CFH* p.Pro503Ala. In our larger present study, *CFH* ENSP00000356399.4:p.Pro503Ala was automatically analysed under both autosomal dominant (AD) and autosomal recessive (AR) models for primary immunodeficiency diseases. The updated scoring methods rank this with a score of 4. For example, it is flagged as benign on ClinVar and a more accurate MAF (0.003943) is found with a newer gnomAD GRCh38 database. Our revised scoring methods assigned a pathogenicity max score of 4 under the more lenient AD criteria, falling below the cutoff of 6 required for inclusion in the main results of our study. Although this score does not support prominent reporting in the larger cohort, it does not negate the findings of the earlier report and remains relevant for individual case analysis.

Similarly, the variants *CYBB* ENSP00000367851.4:p.Gly364Arg and *WAS* ENSP00000365891.4:p.Glu131Lys, associated with X-linked diseases, were reevaluated. While these variants are clinically relevant for individual patients, they did not achieve high enough scores to warrant prominent reporting in our expanded cohort study. Specifically, the *CYBB* variant was identified in 5 individuals, with one male exhibiting a hemizygous genotype. The WAS variant was found in 4 individuals, with one also displaying a hemizygous genotype. Neither variant met the strict pathogenic criteria set by ACMG guidelines, with the ClinVar database listing them as likely benign, thus affecting their evidence score.

This highlights the critical importance of continuous and comprehensive evaluation of genetic variants against updated criteria and multiple reference databases to avoid overly reliant conclusions on any single source. Although not ranked highly enough for main text inclusion, these variants are still considered justifiably reported as candidate causal for the individual patients.

We report the entire dataset as supplemental table “ACMGuru_singlecase_genetic_df_report.csv” which has 2325 qualifying variants regardless of their final score. Previously reported variants are listed but prioritised lower due to their relative significance in the larger cohort. It is confirmed that no alternative variants for these three patients ranked higher in our findings.

Borghesi et al. 2020 also reported 41 additional supplemental VUS (Borghesi Supplementary_Table_4.tsv). We wanted to see if these were still present in the input dataset. Previously, Borghesi et al. 2020 used reference genome GRCh37 and reported only cDNA or amino acid positions without transcript information or genomic coordinates, there the genomic coordinates were used. The genomic coordinate positions were formatted with the addition of “chr” and +1 to create a bed file which was converted with UCSC hgLiftOver (https://genome.ucsc.edu/cgi-bin/hgLiftOver) to update the genomic coordinates from GRCh37 to GRCh38. We include these datasets in the analysis code repository.

This contained 1024 variants in 303 genes, for all variants with MAF <0.1 and which overlap with the PID gene list at that time. In our cohort, after filtering for qualifying variants from GATK4, we had a total of 1257 matches (multiple carriers in some variants), with 459 unique variants. After scoring with ACMGuru we confirmed why these variants were not reported as top candidates in our expanded cohort. There were 273 variants with an evidence score >0, of which 35 were unique variants. Of the variants that pass QV filters, 17 are reported in our unfiltered supplemental table with new evidence scores: *BACH2* p.Glu283Lys, *CFH* p.Pro503Ala, *CHD7* p.Ala99Val, *CHD7* p.Pro101Leu, *CHD7* p.Pro2351Leu, *CHD7* p.Pro900Arg, *CYBB* p.Gly364Arg, *IL17F* p.Arg131Trp, *IRAK1* p.Arg366Cys, *KMT2D* p.Arg2905His, *NFAT5* p.Gly389Ser, *POLA1* p.Asp1208His, *PTEN* p.Ser360Gly, *SAMD9* p.Arg1040Cys, *SEMA3E* p.Pro171Ser, *TNFRSF13B* p.Glu140Lys, *WAS* p.Glu131Lys. The remaining variants did not pass upstream qualifying variant thresholds and thus have no evidence scores for inclusion in our supplemental tables.

**Figure 13** (validation of previous study) shows the raw count of variants detected in the previous study matched to our study and applies the ACMGuru evidence scoring method. Interestingly, we see that it is now possible to classify variants quantitatively as candidate causal due to the updated criteria and evidence sources. Importantly, some of these scores are *artificially inflated by necessity* since we had to remove the qualifying variant frequency filters which otherwise eliminate the variant during processing. We did not record any of these variants passing the reporting threshold. This reinforces the results of the previously study in reporting these as VUS.

Additionally, in our analysis of the three patients highlighted in Borghesi et al. (2020), we did not identify any new leading candidate variants beyond those previously reported. Although we discovered 29 additional qualifying variants, they did not achieve the evidence scores required for higher-priority reporting and were classified as variants of unknown significance (VUS).

This type analysis also raises a point worth consideration for future studies. It is possible to encounter a patient that has conflicting results. It is relatively common to encounter variants that are ranked second which are intuitively the top candidate to experienced reader. For example, in our study, we manually re-checked for X-linked diseases in case there was any erroneous prioritization. We had 42 cases where an X-linked disease gene was detected with genotype 2 (compound het, homozygous, hemizygous). Of these, seven had scores >=6 and all of these were automatically reported in our main text table 1. Therefore, while in our study these were correctly reported as top candidates for the patient, it might not always be so, illustrating that challenging caveats will remain.

Our analysis passes the first challenge of automated evidence quantification and prioritizes interpretation of variants that pass the reporting threshold. The remaining hurdle is the countless nuanced criteria to prioritize disease-specific priors in order to make the final determination. Today, this still often depends on the researchers “intuition”. Even commercially, classification software generally provides automation of this final result for “research use only”. We continue to improve such downstream progress for future work (e.g. https://github.com/DylanLawless/heracles is currently under development). It is likely that the biggest breakthroughs will come by the application of ML/AI, DL/RL, or Bayesian frameworks to provide automated interpretation supported by confidence metrics

#### 1.1.3 Conclusion

Our reevaluation of rare variants in primary immunodeficiency genes from the Borghesi et al. (2020) study not only re-identified the three top candidate causal variants reported in their main results but also reaffirmed their independent classification as the top candidates. We confirmed the presence of 1024 variants originally screened in the smaller cohort, including those listed in the supplemental VUS report. However, despite employing updated and expanded evidence sources and stricter scoring methods, we found no additional evidence to classify any of the VUS as pathogenic.

## 2 Supplemental details

### 2.1 Result note: main analysis features

In our statistical genomics study, the primary analyses included single variant association tests (typical of GWAS), single gene, and protein pathway variant set association tests (VSAT). The protocols for GWAS are well-established, and the VSAT protocol follows a similar data preprocessing approach. For single gene VSATs, the variant set ID encompasses all variants from a specified gene. These variants are then retrieved, and the test is performed on this subset. For pathway-level VSATs, the variant set comprises genes distributed across the genome. The resulting test statistic is reported with the variant set ID (or pathway ID), along with associated metrics such as the p-value, list of involved genes, and variant counts. For instance, the final pathway printed to logs during automated analysis had p-value of 0.187, with 8 genes, 25 unique variants, and 23 and 9 heterozygous variants in cases and control, respectively. To maintain modularity and manage data size, we stored data in .vcf.gz and .vcf.gz.tbi files, split by chromosome, and imported variants only when needed for gene or protein pathway analysis and interpretation.

For our top association pathway, the dataset imported by ACMGuru had 867 variants, some of which were shared in multiple samples, and a final total of 355 unique variants. This is accompanied by 300 columns of annotation information, approximately 10 of which are specific to the sample (sample ID, quality, etc.). Therefore, the final annotation and interpretation table for ACMGuru is a relatively small matrix (867 × 300) suitable for desktop analysis in R, python, etc.

For single variant level analysis we used plink v1.9 to convert from gVCF to .bim, .bed, .fam format. We used the same input (as SkatRbrain for VSAT analysis) of 201874 variants with a genotyping rate of 0.89. We applied a missingness filter --geno 0.05 with 125975 variants were retained by plink for association testing. Any variants with genotype missingess were similarly filtered in VSATs at the individual chromosome VCF level.

### 2.2 Result note: automated clinical cohort summary

ACMGuru was automated to perform a cohort summary for single case analysis using the study handbook variables. Variants which were considered pathogenic or likely pathogenic (ACMG total score = 6) were retained. After analysis, 64 patients were identified. Values are indicated as median [min - max].

Demographic Information - Number of previous sepsis episodes registered in the same child (episode.nr.stats): 1 [1 - 1]. Patient age at blood culture sampling in days (age.at.bc.days.stats): 295 [0 - 5430]. Gender distribution of the cohort (sex.distribution): female: 25, male: 40. Age categories based on the time of blood culture sampling (age.grp.distribution): child.10y.16y: 6, child.1y.4y: 17, child.5y.9y: 8, child.less12mt: 10, neo.preterm: 14, neo.term: 10. Hospitalisation Data - Hospital stay length after blood culture sampling in days (hosp.los.bc.stats): 16 [0 - 230]. Total length of stay in the PICU in days (picu.los.stats): 17 [1 - 269]. PICU stay length after blood culture sampling in days (picu.los.bc.stats): 11 [1 - 216]. Delay in hospital admission from time of initial presentation in days (hosp.delay.stats): 0 [-2 - 202].

Clinical Outcomes - Total number of organ failures as defined by the 2005 consensus (cons05.score.agg.stats): 1 [0 - 5]. Total number of organ failures as defined by the PELOD- 2 (pelod.score.agg.stats): 2 [0 - 23]. Total number of organ failures as defined by the 2017 pSOFA (psofa.score.agg.stats): 4 [0 - 21]. Mortality outcomes within 30 days post-admission (death.30.bc.distribution): no: 64, yes: 1. Impact of PICU length of stay on outcomes (picu.los3.distribution): no: 37, yes: 28.

Organ Failures and Sepsis Details - Cardiovascular failure score under 2005 consensus definitions (cons05.cvs.agg.distribution): no: 56, yes: 9. Respiratory failure score under 2005 consensus definitions (cons05.resp.agg.distribution): no: 40, yes: 25. Central nervous system failure score under 2005 consensus definitions (cons05.cns.agg.distribution): no: 60, yes: 5. Renal failure score under 2005 consensus definitions (cons05.ren.agg.distribution): no: 59, yes: 6. Hepatic failure score under 2005 consensus definitions (cons05.hep.agg.distribution): no: 60, yes: 5. Haematological failure score under 2005 consensus definitions (cons05.hem.agg.distribution): no: 49, yes: 16. PODIUM score (podium.score.agg.stats): 1 [0- 8].

Pathogen Information - Primary clinical focus or reasons for medical intervention (focus.grp.distribution): abdominal: 4, clabsi: 14, cns: 3, earnosethroat: 2, osteoarticular: 8, other: 2, pneumonia: 3, primbsi: 14, skin: 4, uti: 9, wound: 2. Types of pathogens identified in blood cultures (pathogen.grp.distribution): candida: 2, cons: 9, ecoli: 10, enterococcus: 1, hinfluenzae: 3, klebsiella: 5, othergneg: 10, paeruginosa: 4, sagalactiae: 3, saureus: 8, spneumoniae: 3, spyogenes: 5, viridansgroup: 1.

### 2.3 Result note: single-case analysis

We performed statistical analysis to see if any clinical features were significantly associated with variant consequences found in single-case analysis. The negative result was included since any clinical feature that is a strong marker of genotype/phenotype would provide insight for future cases and would be of importance for downstream interpretation, and it is thus worth noting this test outcome. Since our cohort, by design, consisted of patients grouped with suspected rare disease there was no assumption that such as association would or would not exist.

### 2.4 Result note: Single variant analysis

For single variant analysis in Archipelago plot we used 1df chi-square allelic test. More complex models were also applied for single-variants tests; however, we show the simplest method for clarity since we had no single-variant enrichment.

### 2.5 Result note: Protein pathway construction

No additional discussion.

### 2.6 Result note: Interpretation of VSAT

The frequency of individual variants which are present within the enriched pathway (variant set ID 22 / 586) are illustrated in **figure 4** (A). **Figure 4** (B) illustrates the number of unique variants per gene in pathway ID 22 / 586. The contributions of low and high frequency variants are expected to differ. Therefore, it is important to note that the statistical analysis using our VSAT protocol (SKAT-O) accounts for the MAF and therefore variants which are present at higher frequencies, such as common and homozygous, are naturally down-weighted automatically thereby accounting for their potential small effect size. In traditional single-variant analysis such as those using simple logistic regression or Fisher’s test, such variants might be better removed from the QV set for rare disease analysis since they are unlikely to contribute strong effect for individuals. However, with combined burden and variance component tests we can thus retain all QV for improved detection and reduce potential for type II error.

## 3 Supplemental extended methods

### 3.1 Sub-setting and variant collapse

Our data was prepared in cohort batches with all cases and controls. Joint genotyping and downstream analysis with GATK best-practices was applied to the joint cohort. Statistical analysis required modifications from the joint cohort dataset depending the computational resource requirements, as follows.

Single-case analysis consisted of annotating and interpret all variants in the joint dataset together and only in the final stages, we reported variant carriers where a variant passed our filtering criteria. Therefore we jointly analysed the fully annotated dataset (GATK to VEP with all default and custom plugins). ACMGuru was applied to retrieve final variant results.

Gene-level testing required us to perform statistical analysis on a large dataset in parallel. Therefore, we output a second version of VEP annotation using a minimal set of plugins to reduce non-essential data stored in the CSQ info field of VCF. This allowed us to retain the same filtering information while greatly reducing the memory requirements. Variant collapse was performed at the gene-level using SKATRbrain. ACMGuru was to be applied to retrieve final variant results, however since no single genes were significantly enriched this step was omitted.

Pathway-level testing was performed on the same data as the gene-level description. Variant collapse was performed at the pathway-level using SKATRbrain. ACMGuru was applied to retrieve final variant results on significantly enrich pathways.

Single variant testing was performed on the same data as gene-level and pathway-level data, however, plink v1.9 was used to convert from VCF to bed, bim, fam format. We used plink v1.9 to perform single variant association testing using --assoc and other modes. ACMGuru was applied to retrieve final variant results for significantly enriched variants. The relevant details from each output were passed to Archipelago (methods) to perform the archipelago summary (p-value, study type, gene symbol, pathway (variant set ID), chromosome, position, and variant ID). Covariates used in statistical analysis were the same throughout, all derived from the upstream joint dataset.

Since we first cluster the genome-wide protein pathway data independently, variant collapse was simply performed by grouping variants based on the gene or pathway ID in each of the relevant tests. Candidate variants remaining after filtering based on the criteria in section 7 were collapsed into genes and/or protein pathways. An overview of methods for variant collapse is reviewed by Povysil et al.

Correction for multiple was testing based on the total number of tests was used in each study and is indicted by the significance thresholds throughout. In general, approximately 19000 protein coding genes are included in exome sequencing studies, and therefore a significance threshold relative to the number of tests is used; for gene-level testing this is approximately *α* = (0.05*/*19000) ≈ 2.6*x*10^−6^ (Povysil et al.). In protein pathway analysis approximately 1000 pathways were present, resulting in a pathway-level threshold of approximately *α* = (0.05*/*1000) ≈ 5*x*10^−5^. Exact values are shown where used based on the number of tests run by SKAT-O or plink.

### 3.2 Protein pathway construction overview

A protein pathway interaction (PPI) network was generated to determine a “best fit” for all proteins genome-wide based on independent reference database before applying it to the cohort dataset. With this new reference genome-wide PPI database, we can compare future studies as well as removing potential bias from restricting pathway definitions to those which suit our dataset. Details and an R package for performing this step, ProteoMCLustR, is described in the next section. Briefly we discuss the overview. Protein interaction data was sourced from several databases as compiled by STRING database (Szklarczyk et al. 2016). The STRING db is a comprehensive resource for PPI, which integrates both curated data sources - GO, KEGG, Reactome, Biocarta, and BioCyc, and experimental sources - BIND, DIP, GRID, HPRD, IntAct, MINT, and PID.

With ProteoMCLustR, we downloaded the STRINGdb, species 9606, version 11.5 (Szklarczyk et al. 2016). This consisted of 19,566 proteins and the number of known interactions varied according to confidence levels: 1,795,134 low; 505,968 moderate; 247,200 high. These STRING db confidence score cutoffs were considered when constructing our final protein pathway reference dataset. We tested at the three recommended levels - 0.4, 0.7, and 0.9 (otherwise labelled as 400, 700, 900, respectively within the dataset). This confidence score is the measure of evidence required to create an interaction edge (PPI) between two nodes (proteins).

Iterative reclustering was run as part of MCL by ProteoMCLustR with parallelisation to reduce the time required for genome-wide clustering to *<* 10 hours per confidence score dataset. This process tested multiple network inflation parameters and numerous iterations (defaults for human genome in ProteoMCLustR) to find the configuration under which proteins belong to a pathway. We ran the same network construction protocol on each set. The output after clustering was an array of individual protein pathways consisting of gene symbols and a pathway id (MLC group ID of the variant set based on order of completion). The STRINGdb confidence score 700 resulted the largest number of retained proteins assigned to pathways, thus we continued with this network of PPI as the optimal balance between evidence reliability and retention of proteins compared to the maximum cut-off (900) which results in a large loss of proteins as singletons. Note that singletons which have no PPI are dropped during clustering, and a cut-off pathway size of minimum 5 and maximum 50 is used to reduce pathways to biologically interpretable sizes (details in further section). These prepared genome-wide PPI datasets are available in the ProteoMCLustR package for reuse.

### 3.3 ProteoMCLustR

In this study, we designed and used the ProteoMCLustR method which combines the Markov cluster algorithm (MCL), STRINGdb (or any alternative PPI database), and customizable parameters tailored to the biological scenario (such as PPI evidence score, and biological pathway size limits) (Szklarczyk et al. 2016, Van Dongen SM. 2020). Construction of whole-genome protein pathways was based on these external databases independently of the association test data. It is therefore less prone to bias during the subsequent statistical analysis. For context, the subsequent test (expanded in further sections) entails of collapsing the cohort genotype into one variant set per protein pathway - as produced by ProteoMCLustR - and then performing case-control analysis by VSAT using SKAT-O, thus analysing every protein pathway, genome-wide. This approach enables a comprehensive understanding of the biological networks while considering their complexity and specificity.

The protocol is summarised in **Figure S5** and detailed steps are further provided in the extended supplemental methods.

### 3.4 Markov Cluster Algorithm (MCL)

Step 1: Input Graph

- Begin with an input graph represented as an adjacency matrix or a weighted graph.

Step 2: Expansion

Start with an initial matrix, often called the *stochastic matrix*, denoted as *M*(^0^), where each element represents the probability of transitioning from one node to another in a random walk.

Raise the matrix to a power, typically denoted as *e*, to simulate multiple steps of the random walk:

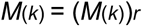

This step helps in emphasizing highly connected regions in the graph.

Step 3: Inflation

- Apply an inflation operation to the matrix obtained from the expansion step.
- The inflation operator consists of two main operations: expansion and renormalization.

Expansion:

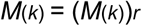

where *r* is a positive power (usually greater than 1), which accentuates strong connections and suppresses weak connections.

- Renormalization:

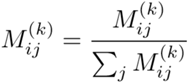

This step ensures that the matrix remains stochastic, i.e., each column sums up to 1. Step 4: Repeat Expansion and Inflation

Repeat the expansion and inflation steps iteratively until convergence is achieved.

Convergence can be determined by measuring the change in the matrix between iterations or by reaching a specified number of iterations.

Step 5: Cluster Identification

Identify clusters in the final matrix, denoted as *M*^(*final*)^.

This can be done by applying a threshold to the elements of the matrix to distinguish between cluster and non-cluster elements.

The resulting clusters can be represented as sets of nodes or as separate subgraphs.

Step 6: Post-processing

Perform any desired post-processing steps on the identified clusters.

Examples of post-processing include merging similar clusters, filtering out small clusters, or assigning cluster labels based on the nodes’ attributes.

Step 7: Output

Output the final clusters obtained from the MCL algorithm as the result.

### 3.5 ProteoMCLustR protocol

1. Set the ‘file suffix’ variable and redirect R output to a log file using the ‘sink’ function.
2. Load required packages, including ‘STRINGdb’, ‘parallel’, ‘dplyr’, and ‘ggplot2’.
3. Instantiate a ‘STRINGdb’ object by connecting to the STRING database (version 11.5) with a specified score threshold and save the data as an RDS file.
4. Read protein information from the file, selecting the relevant STRING ID and Gene ID columns, to create a data frame called ‘string id_df’.
5. Define the ‘GetSubNetwork’ function, which iterates over a set of protein IDs to extract a subnetwork from the STRING database.
6. Define the ‘ChooseInflation’ function, which iterates over a set of MCL inflation parameters (‘mcl_inflation_param’) to find the optimal parameter for clustering a given subnetwork. The function utilizes the ‘cluster markov’ function from the SoloTonic package and runs MCL clustering in parallel for different inflation parameters.
7. Define the ‘RunMCL’ function, which recursively runs MCL clustering on a subnetwork using the ‘ChooseInflation’ function. The recursion continues until a certain iteration limit (‘iter_limit’) or cluster size limit (‘size_limits’) is reached. If a cluster exceeds the size limit, the function recursively calls itself on the overlimit clusters.
8. Run the ‘RunMCL’ function on the whole genome using the ‘string id df’ data frame.
9. Recode the STRING IDs to gene IDs, creating ‘mcl clusters recode’, and save the clustering results to RDS files.
10. Generate a data frame from the clustering results, with each row representing a gene and its corresponding pathway ID.
11. Group the rows by pathway ID and count the number of genes in each pathway. Plot the counts using ggplot2 and save the plots as PDF files.
12. Check for overlaps between the groups by generating all possible combinations of two groups, counting the number of shared genes in each pair, and creating a data frame with overlap counts.

The MCL algorithm is a graph clustering technique that identifies clusters in an input matrix by iteratively applying expansion and inflation steps until the matrix converges to a stable state. The custom ‘RunMCL’ function uses an ‘iter limit’ parameter to control the depth of recursive calls and ‘size_limits’ to manage cluster size. The function recursively calls itself on subnetworks corresponding to clusters exceeding the specified ‘size limits’ until no cluster exceeds the size limit or the ‘iter limit’ is reached. The ‘ChooseInflation’ function determines the optimal MCL inflation parameter, affecting the granularity of the clustering output, and is used in the main ‘RunMCL’ function.

The MCL algorithm works on the entire network simultaneously, refining its structure until distinct clusters emerge and the entire network reaches a stable state. The custom script checks the size of the resulting clusters after the MCL clustering is complete. If a cluster exceeds the specified ‘size limits’, the script will re-run the MCL algorithm on that specific large cluster with an adjusted inflation parameter, recursively until clusters fall within the defined ‘size limits’ or the maximum allowed iterations (‘iter limit’) is reached.

The algorithmic description is as follows:

Input:

*N_i_, i* = 1*,…,n* : Nodes (genes) in the STRING database

*E_ij_, i,j* = 1*,…,n* : Edges (interactions) between nodes in STRING database

*S* : Score threshold for edges

*I* : Iteration limit

*L*_min_*,L*_max_ : Size limits for clusters

*e,r* : Expansion and inflation parameters for MCL algorithm

Algorithm:

[1] Preprocess 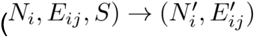
[2] ChooseInflation 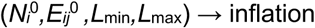
[3] RunMCL 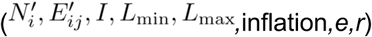
[3.1] Initialize 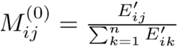
[3.2] Iterate until convergence:
[3.2.1] Expansion 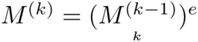
[3.2.2] Inflation 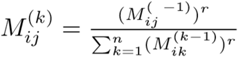
[3.3] Extract clusters from converged matrix *M*^(final)^

Output:

Clusters: Set of optimized node (gene) cluster where *k* is the index variable used in the iterative steps of the MCL algorithm, representing the iteration number. Node indices in the graph are represented by *i*, *j*. In the context of the adjacency matrix or the stochastic matrix, *i* represents the row index and *j* represents the column index. *M*^(*k*)^*ij* represents the element of the stochastic matrix *M*^(*k*)^ at the *i*-th row and *j*-th column, representing the probability of transitioning from node *i* to node *j* during a random walk on the graph at the *k*-th iteration of the MCL algorithm.

*M*(^0^)*ij* represents the initial stochastic matrix.

### 3.6 MCL parameters

ProteoMCLustR was used to apply MCL. An alternative graphical user interface approach would be possible with the Cytoscape application (Shannon 2003) using the clustermaker 2 plugin for Cytoscape although the memory, time, and variable management may be difficult, therefore we recommend our R package on a HPC environment for whole-genome level data.

For our inflation parameters we set seven intervals: 1.5, 2, 2.5, 5, 10, 15, 20. Our number of interactions were set to 4. The network data was exported in both CSV and Rds format, consisting of gene symbol and MLC group ID (pathway ID based on order of completion).

### 3.7 Pathway size threshold

Since our study type was based on suspected rare disease, we set our criteria for thresholds to best reflect the likely biological scenario. For example, protein pathways which have greater than approximately 50 proteins would likely have redundancy that outweighs the effects of single variants in a single individual. This value is likely to be variable depending on the phenotype and expected effect size detectable. Likewise, pathways which are too small (i.e. *<* 5 proteins) lack sufficient interaction information for biological or statistical interpretation.

Therefore, we set these limits as our maximum and minimum pathway size for clustering. However, we also note that if an association present in larger networks (i.e. *>* 50) it would also likely be detected in each of the smaller subsets when sample size is sufficient.

### 3.8 VSAT significance testing

SKAT and its optimal unified version SKAT-O are now popular methods for gene-based association tests, accommodating multiple variants within a gene or variant set while accounting for their potentially differing directions and magnitudes of effects. Here, we briefly expound upon SKAT and SKAT-O tests, as well as the resampling method typically employed for calibrating these tests, the methods as used in our analysis. We previously also compared a range of association testing methods on synthetic data to assess performance of methods including simple logistic regression, Fisher exact test, logistic regression, CMC, weighted-sum test, SKAT, SKAT-O, etc. Our testing agreed with the recommendations previously reported, finding the SKAT-O methods best suited for this disease study type as demonstrated by Lee et al. (2012). SKAT-O is based on the original implementation of SKAT by Wu et al. (2011), and demonstrated by Lee et al. (2012) and Ionita-Laza et al. (2013). Nicolae (2016) has reviewed many of the additional similar methods, although none were more appropriate for our usage. We also applied the resampling method for calibrating single and gene-based rare variant association analysis in case–control studies by Lee et al. (2016)

SKAT and SKAT-O are both powerful methods used in gene-association studies, particularly when considering rare genetic variants. SKAT uses a kernel-based approach, while SKAT-O uses a combination of burden tests and SKAT. For SKAT, the dataset consists of *n* subjects sequenced in a region (gene or variant set) with *m* variants. Let *y_i_* denote a dichotomous phenotype, *G_i_* = (*g_i_*_1_*,…,g_im_*)^0^ the genotypes of the *m* variants (*g_ij_* = 0,1,2), and *X_i_* = (*x_i_*_1_*,…,x_is_*)^0^ the covariates.

In the logistic regression model: 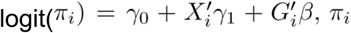 is the disease probability, *γ*_1_ isan *s* × 1 vector of regression coefficients of covariates, and *β* = (*β*_1_*,…,β_m_*)^0^ is an *m* × 1 vector of regression coefficients of genetic variants.

The SKAT statistic is then typically written as: *Q_S_* = (*y* − *π*^)^0^*K*(*y* − *π*^), where *π*^ = (*π*^_1_*,…,π*^*_n_*)^0^is a vector of the estimated probability of *y* under the null model, and *K* = *GWWG*^0^ is an *n* × *n* kernel matrix, where *G* = (*G*_1_*,…,G_n_*)^0^ is an *n* × *m* genotype matrix, and *W* = diag(*w*_1_*,…,w_m_*) is an *m* × *m* diagonal weight matrix.

SKAT-O combines the advantages of the burden test and SKAT into a single unified test. The statistic for SKAT-O is: *Q_ρ_* = *ρQ_B_* + (1 − *ρ*)*Q_S_,* 0 ≤ *ρ* ≤ 1, which is a weighted average of the SKAT and burden-test statistics. The optimal weight *ρ* is estimated from the data to maximize the test power. The optimal unified test is then: *Q*_optimal_ = min_0≤*ρ*≤1_ *p_ρ_*, where *p_ρ_* is the *p*-value computed on the basis of a given *ρ*. For large samples, each test statistic *Q_ρ_* can be decomposed into a mixture of two random variables; one asymptotically follows a chi-square distribution with one degree of freedom, and the other can be asymptotically approximated to a mixture of chi-square distributions with a proper adjustment. Under the null hypothesis, the statistic *Q* follows a mixture of chi-square distributions. Its p-value can be calculated using Davies’ method or a fast moment matching algorithm (Davies 1977, Davies 1980, Davies 1987, Davies 2002). Hence, the *p*-value of *Q*_optimal_ can be quickly obtained analytically with the use of a one-dimensional numerical integration. Small sample sizes (*<* 2000) adjustments are applied accordingly as part of the SKAT R package for our cohort (n = 940).

To calibrate the tests and adjust the Type I error rate, the SKAT and SKAT-O methods often employ a resampling procedure. The method by Lee et al. (Lee et al. 2016) is implemented in the R package SKAT, which we used for SkatRbrain. This involved generating permuted datasets by randomizing the phenotypes, and then recalculating the test statistics for each permuted dataset. The p-value of the observed test statistic is then calculated as the proportion of permuted datasets for which the permuted test statistic exceeds the observed one. This method can be computationally intensive and can also be approximated by more efficient techniques, such as the parametric bootstrap method. We apply variables in SkatRbrain (described in the next section) to use a default or custom resampling value for a user selection of variant set subsets since large datasets can be time consuming and only a small number of sets may need reinvestigation.

We hypothesised that none, or a small level of enrichment, should be seen for single variants due to the necessity to include only rare variants. For the same reason, we expected few or no enriched genes or protein-pathways, especially when resampling methods are efficient for controlling type I error (Lee et al. 2016). We expected any significantly enriched signals to be relevant to susceptibility to infection, although no *a priori* requirements, filters, or selections were set. For any significantly enriched signal, our follow-up consisted of annotation-based interpretation.

### 3.9 SkatRbrain

The VSAT process was handled on our high-performance computing environment using our SkatRbrain package. This allows output directly for QV pipeline based on GATK best-practices. Since the large dataset is split into chromosome level files, we provide use SkatRbrain to import each VCF into R. Next, the current protein pathway ID (supplied as output from ProteoMCLustR) is used to make the query variant list. For any variant/gene within this pathway, the individual chromosome VCF are unzipped and filtered iteratively until the full variant set subset is prepared as input for SKAT-O using the SKAT R package. An alternative option is to prepare a plink format subset for every protein pathway ID to pass to SKAT, however we use the former method so that every downstream query on the original GATK VCF output can re-use the same extraction method.

We use vsat-launch to configure and initiate an analysis workflow with customizable parameters. It utilizes HPC environment and is designed to perform either a full analysis or a resampling subset analysis based on the user input.

1. Configuration: The process starts by defining default parameters for the analysis and then allows for these parameters to be altered via command-line arguments. The parameters include a version suffix, total number of samples, number of pathways to test, etc. Different version suffixes correspond to different sets of parameters.
2. Analysis parameters: Depending on the chosen version suffix (v1, v2, v3, v4), various filtering parameters for the analysis are set. These parameters include cohort maximum carriers, rare threshold for gnomAD, and filtering method.
3. Checking for subset resampling: In certain scenarios, additional resampling may be required during set up (Lee et al. 2016). If a file path for resampling a subset is provided when prompted, the script adjusts the parameters accordingly and sets up for a subset resampling analysis. Otherwise, it prepares for a full analysis.
4. Building a submission script: a temporary SLURM job submission script is created automatically which includes the user variables. This script sets up the environment, loads necessary modules, decides the current ID based on whether it’s a subset or full analysis, and calls the main R script ‘vsat-run‘ (for SKAT-O) with the specified parameters for actual analysis. The log and error files from this job are directed to specified directories.
5. Submission: The temporary job script is then submitted to the HPC queue via SLURM.
6. Cleanup: After the submission, the temporary job submission script is copied to logs and removed from the directory.

The output includes result logs, job information, and job logs which are saved in specified directories.

VSAT-run serves as the core of the SkatRbrain method, enabling parallel execution of the VSAT SKAT-O testing on each variant set defined by the variant set IDs (protein pathway IDs). Jobs are launched on the HPC environment such that the scheduler can can assign one job per protein pathway (variant set ID) in parallel. The read parameters include the job ID (variant set ID), input/output sources, the user-defined filter method (i.e. version 1-4), which in turn define the filters such as the gnomAD allele frequency threshold, low genotyping rate threshold, the maximum number of variant carriers in the combined cohort, and the critical number of samples within the cohort. Several subscripts are called for data preparation such as unzipping VCF (.gz), and subsetting the current variant set, genotype cleaning from VCF genotype format to 0/1/2 format genotype matrices for SKAT, a progress bar for monitoring, etc. PCA and covariate data is read in, and variant files are loaded for each chromosome.

Output logs store key analysis results such as the p-value from the SKAT test, pathway ID, analyzed genes, number of chromosomes, and number of samples. Its parallel execution on a HCP cluster makes it highly efficient for large-scale genetic data analysis with run times which match those originally reported by Wu et al. (2011), i.e. approx. 2 hours for approx. 1000 WES samples and testing in 1000 variant sets (protein pathways).

### 3.10 Pathogenicity interpretation

#### 3.10.1 ACMGuru variant classification

Custom methods for gene annotation were used to derive interpretation of any variant which was part of a significantly enriched variant set (single variant, gene, or pathway). VEP was used with all default plugins, as well as custom plugins such as dbNSFP. Scoring was applied to interpret annotation evidence based on the criteria defined by ACMG/AMP guidelines

Richards et al. (27). To do so, we designed and used the ACMGuru R package for automated interpretation and reporting. This process includes summary statistic, text descriptions, and a range of plots describing the interpretation of individual and joint variants. For example, we show the distribution of variants based on several popular pathogenicity prediction metrics, the distribution of variants within genes, pathways, etc. Additional plots are output which illustrate the known information about each gene or protein using data from UniProt and other sources which are mapped onto the gene or protein structure.

The filtering rules for ACMGuru variant-classification follow the ACMG/AMP standard, as per Richards et al. (27) (**Table S2 – S3**). For example the category, evidence of pathogenicity very strong 1 (PVS1) are defined as null variants where the genotype in patiant matches the inheritance type for disease as reported in OMIM, etc, and in gene where LoF cause disease. That is heterozygous null variants in for dominant for known gene/disease, and homozygous or compound heterozygous for recessive disorders. Category evidence of pathogenicity strong 1 (PS1) is defined as the same amino acid change as a previously established pathogenic variant regardless of nucleotide change. We skip some categorisations where they are not appropriate such as PS2 for de novo variants (both maternity and paternity confirmed) in a patient and no family history, since we do not have the possibility of collecting sample for parents. Likewise, we skip category PS4, define as where the prevalence of the variant in affected individuals is significantly increased compared with the prevalence in controls, since we do statistical analysis separately which is more accurate. Some tailored criteria are included as per Richards et al. (27), such as when additional strong pathogenic evidence exists, for example a compound heterozygous variant set with one PSV1 and a second QV in the same gene. This is also similar to pathogenicity moderate 3 (PM3), for recessive disorders detected in trans with a pathogenic variant. PM2 is defined as absent from controls (or at extremely low frequency if recessive) in Exome Sequencing Project, 1000 Genomes Project, or Exome Aggregation Consortium; we do not use “absent in controls” since the upstream QV filtering is stringent and we want to monitor for the presence of any control carriers in the final report. Therefore use gnomAD AF *<* 1*e* − 6 which equals approximately one or fewer heterozygous carriers in gnomAD.

PP3 is defined as multiple lines of computational evidence supporting a deleterious effect on the gene or gene product (conservation, evolutionary, splicing impact, etc.). For predicted pathogenicity interpretation we used VarSome calibrated insilico thresholds from VarSome Germline Classification, version: 11.7.8, 20221202, by Saphetor SA. Their calibrated thresholds were based on the latest ClinVar data, using the ACMG/AMP classifications, using only missense variants, and ignoring: variants with allele frequency greater than 0.01, genes for which no pathogenic missense variants have been reported, entries with 0 stars (literature only, or no approved methodology), entries prior to 2018. We assigned a variant with PP3 when from 22 the engines, there was evidence assigned by ≥ 3 engines as strong pathogenic, ≥ 4 as moderate pathogenic, or ≥ 10 as supporting pathogenic. For each of these three categories this made up less than 1% of the total number of variants in the statistically enriched protein pathway QV and is thus a modest estimate from an already heavily filtered set. **Table S4** lists the exact number of variants on percentage used in each case. We additionally assign PP3 for other commonly used in-silico predictions if ≥ 3 thresholds were passed for: CADD-PHRED ≥ 30, REVEL-rankscore ≥ 0.51, MetaLR “D”, MutationAssessor “H”, SIFT “deleterious”, and PolyPhen “probably damaging”.

## 4 figures

**Figure 1.**
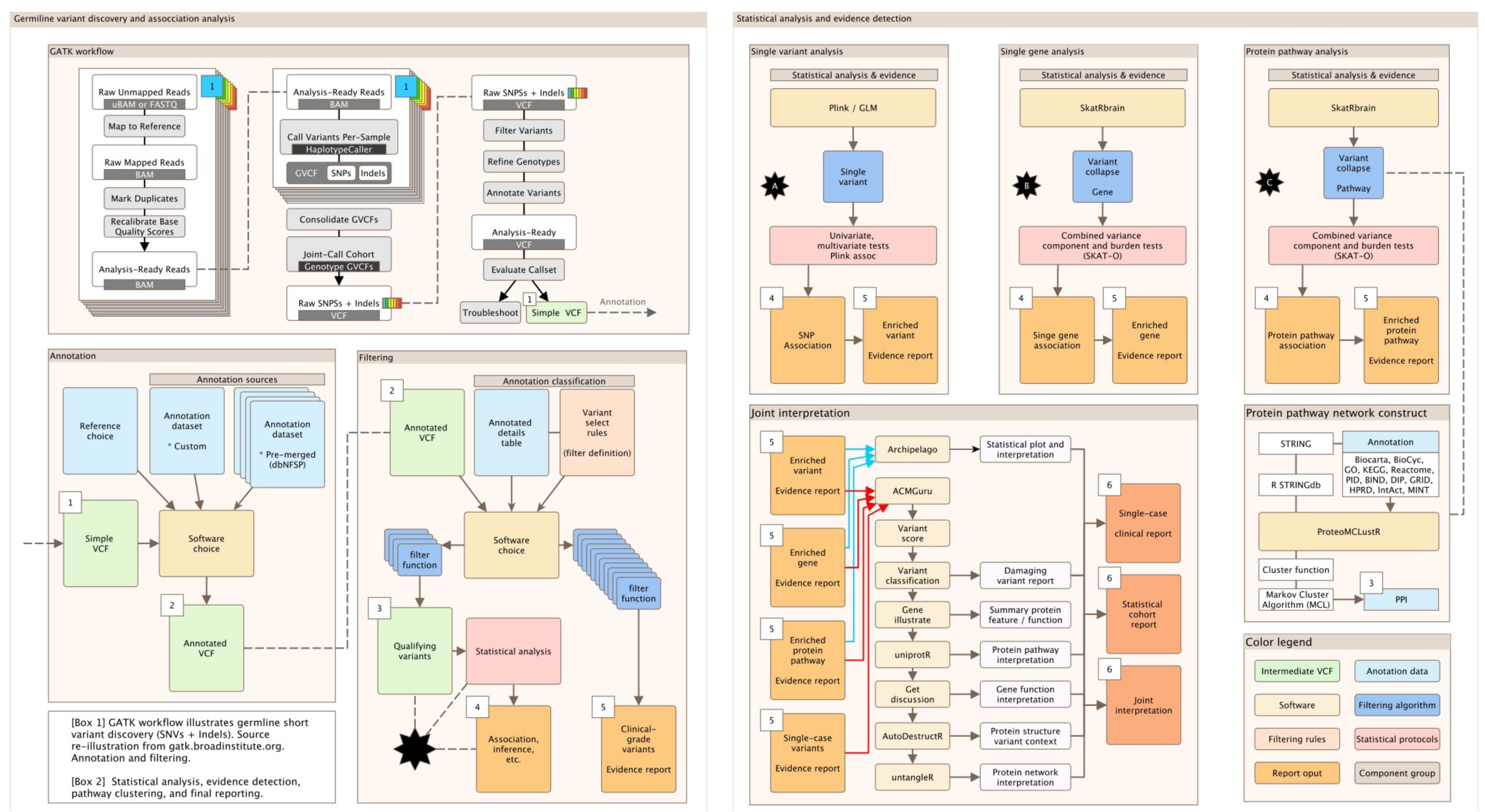
Sequence analysis and association testing workflow detailed. Cohort data consists of exome sequence data, clinical and demographic covariates, and disease status. (Left) Germline short variant discovery which outputs the prepared cohort data VCF (1). Annotation provides rich information about every variant to produce annotated cohort VCF (2). Two filtering options; (i) filtering qualifying variants (3) for statistical testing (4) and (ii) filtering for single-case clinical-type reporting (5). (Right) statistical analysis and interpretation. Statistical analysis of single-variants (A), gene-level VSAT (B), and pathway-level VSAT (C). Each test outputs the summary statistics (4) and supporting evidence (5). A pipeline for interpreting any evidence report (single-case or statistical studies) to summarise all evidence supporting variant pathogenicity, cohort demographics, functional effects, and other explanatory data (6). GLM, generalised linear model; QV, qualifying variant; SVAT, single variant association test; VSAT, variant set association test.

**Figure 2.**
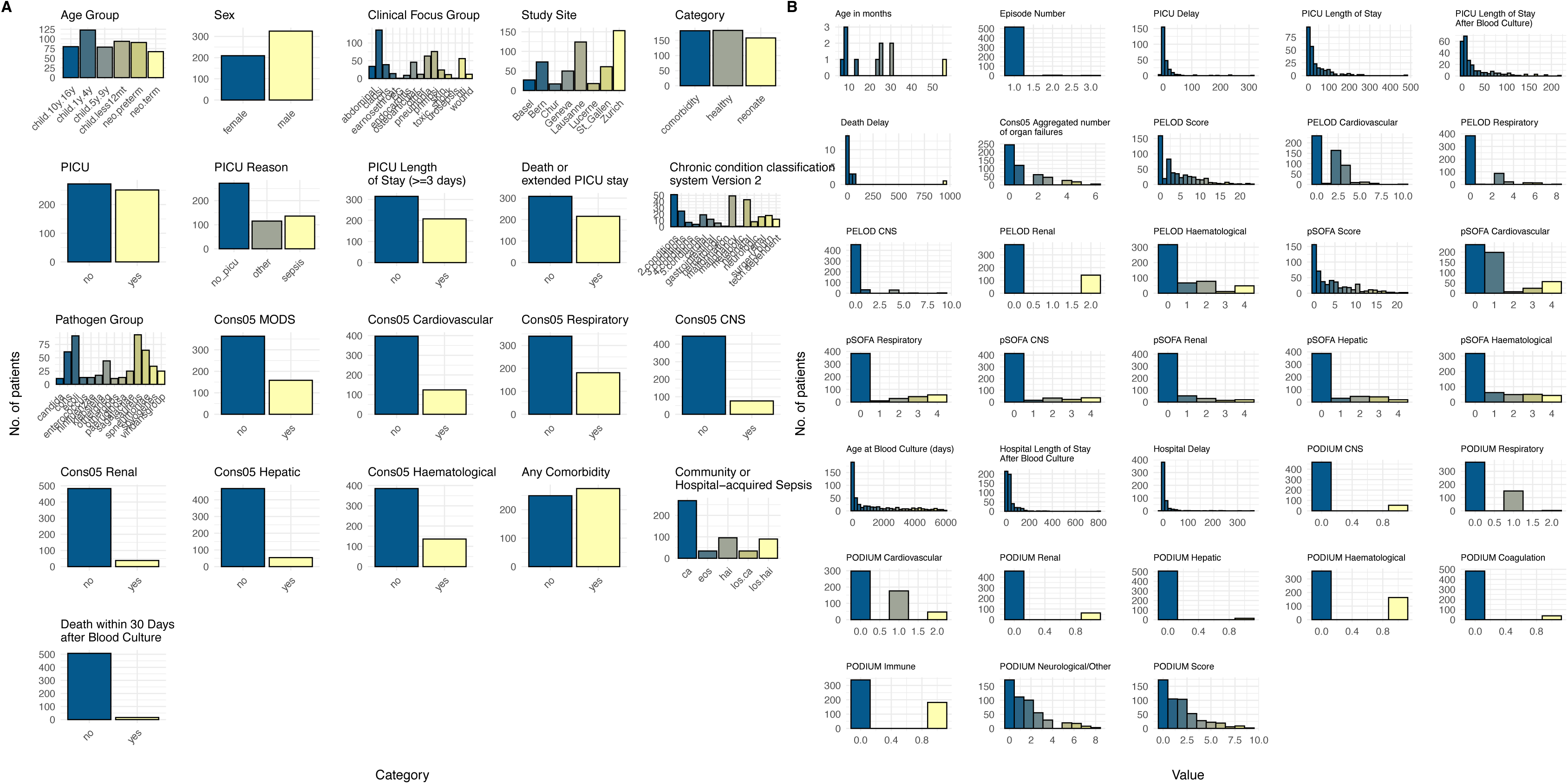
Patient cohort characteristics. (A) categorical and (B) continuous variables for patient cohort. Our cohort consisted of 940 individuals with a total of 548 cases and 392 controls, after QC (quality control) Color scale low (blue) to high (yellow). Focus group labelled EG is ecthyma gangraenosum.

**Figure 3.**
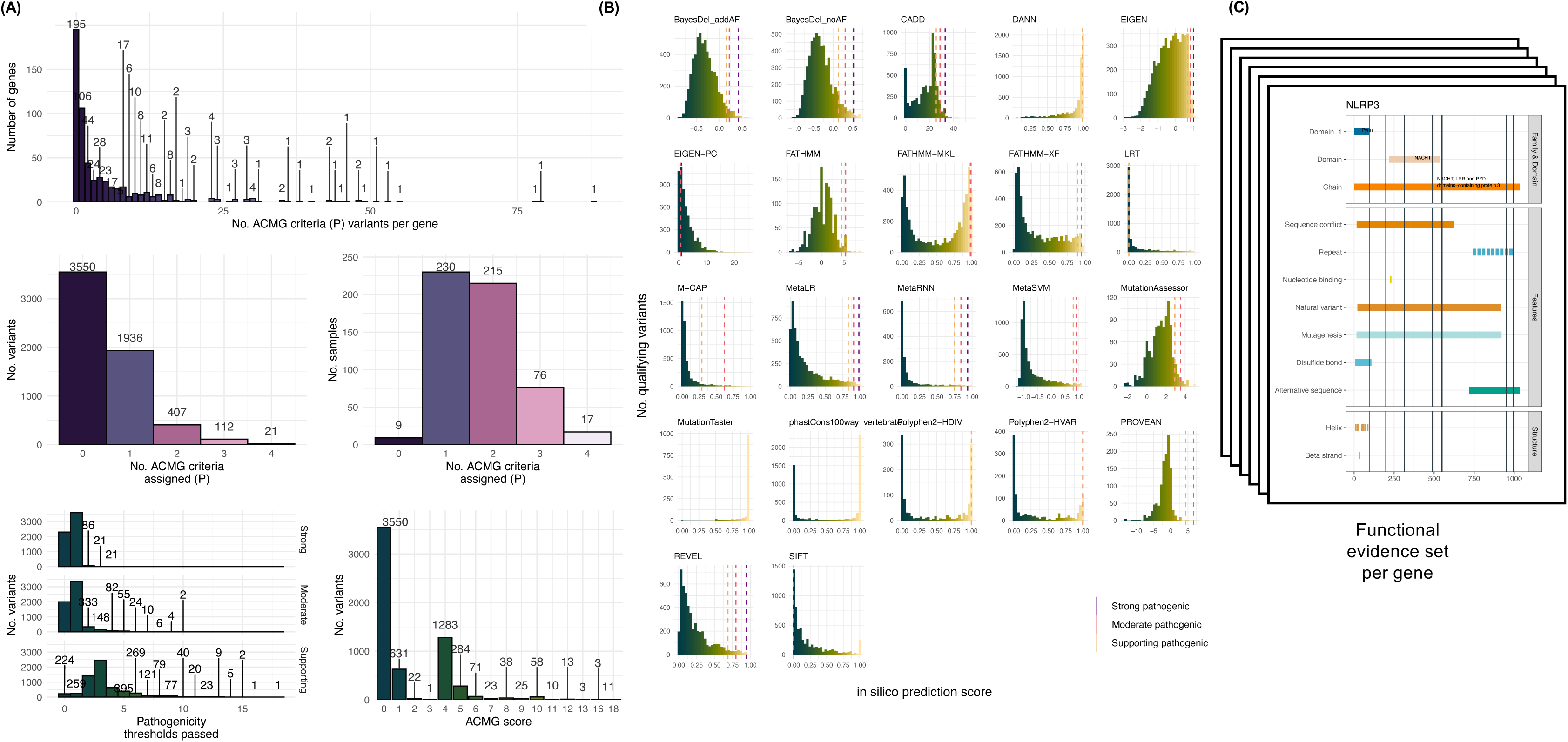
ACMGuru evidence interpretation for single-case analysis of variants. (A) The number of genes versus the number of criteria used for interpreting pathogenicity evidence. (B) The number of unique variants assigned versus the number criteria assigning pathogenicity. (C) The number of samples with unique variants per criteria. (D) Additional variant pathogenicity threshold distribution based on reference sources for strong, moderate, and supporting evidence as seen in (F). (E) Final determination of ACMG/AMP score by ACMGuru. (F) Distribution of variants for each of the prediction engines with reference sources for strong, moderate, and supporting evidence shown by vertical bars. (Bottom) Single-case evidence plots with filter. Genes with at least one variants with ACMG score ≥ 8 (pathogenic) are illustrated. Vertical black colored bars show variants with ACMG score ≥ 8 and other scores > 0 are shown by lighter olive color.

**Figure 4.**
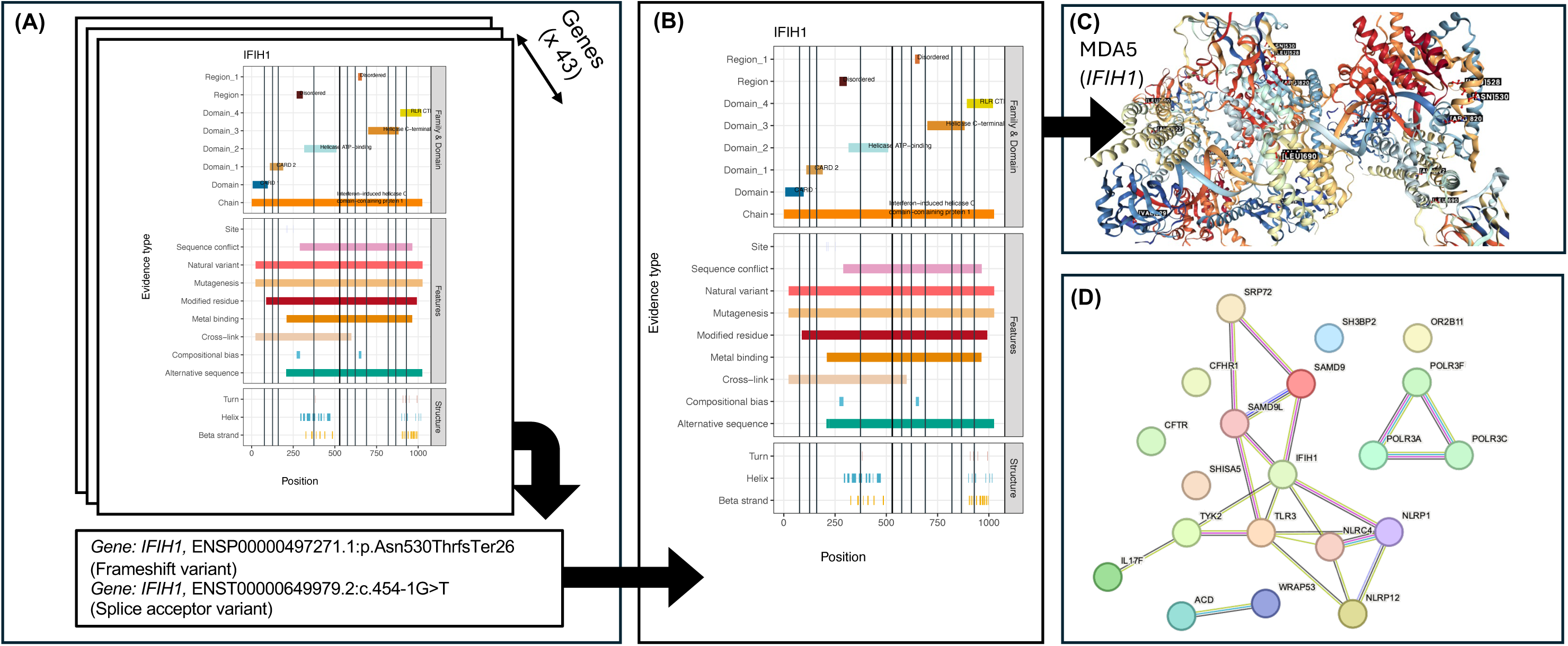
Expanded example ACMGuru interpretation of pathogenicity in single-case analysis. ACMGuru evidence interpretation for single example gene with variants considered as candidate pathogenic for sepsis. We selected this gene arbitrarily based on our background familiarity with its function to assess the automated interpretation. (A) Gene illustrations showing variants considered pathogenic or likely pathogenic. An example of two variants in IFIH1 are highlighted. (B) Gene evidence illustration for the highlighted gene. (C) Protein structure by ACMGuru AutoDestructR shows all variants affecting this protein, used in the interpretation of mechanism. (D) All variants identified as pathogenic or likely pathogenic during single-case analysis were clustered with STRINGdb to provide a overview of related functions for final interpretation.

**Figure 5.**
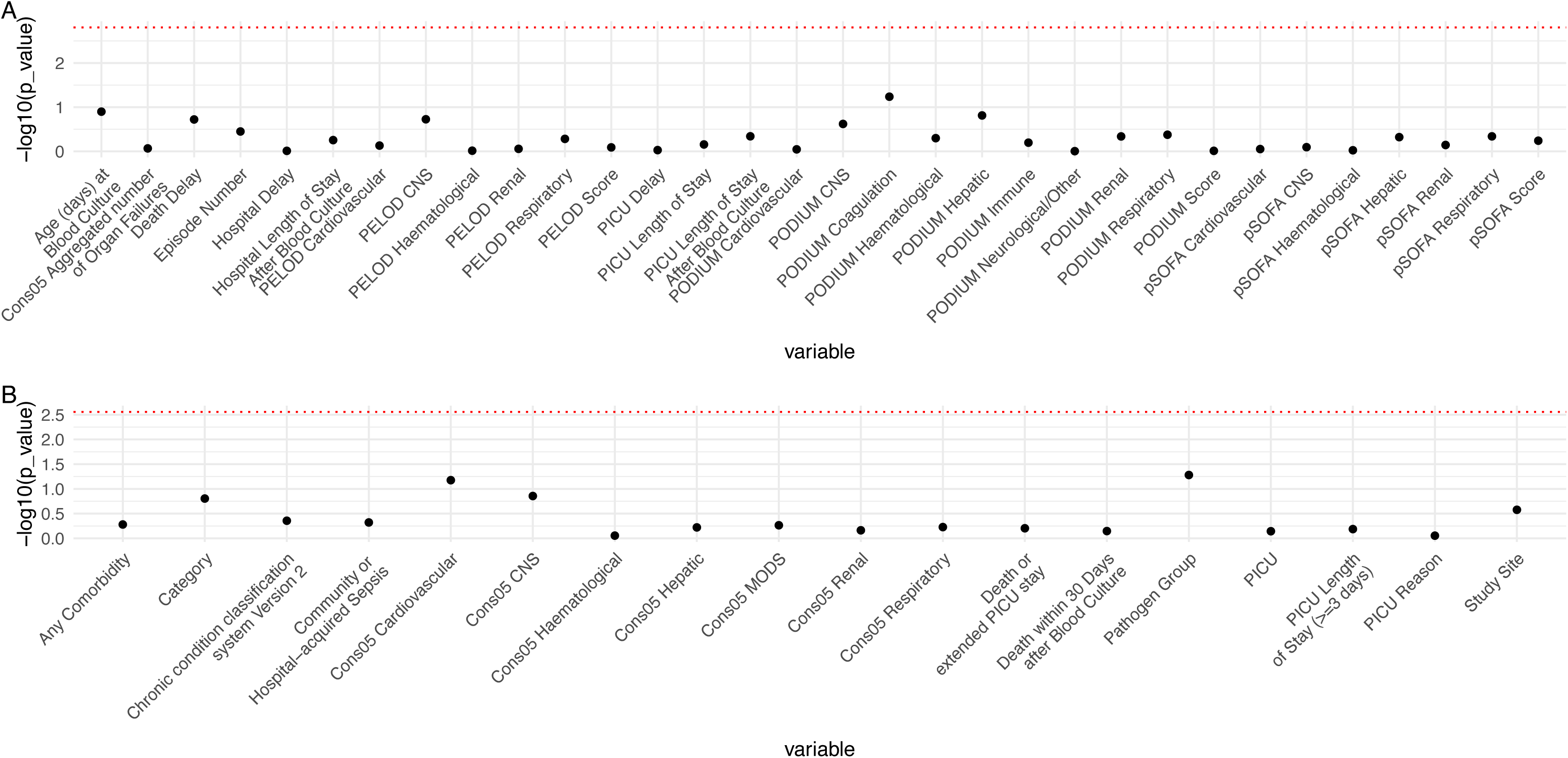
Carriers versus non-carriers of single-case variants with ACMG/AMP score ≥ 4. Any individual with a potentially pathogenic variant with ACMG/AMP score ≥ 4 was labelled. We then performed statistical analysis to see if any clinical features were significantly associated with these variant consequences. Red line indicates the significant threshold after Bonferroni correction. No clinical features were significantly enriched for those with these particular variants.

**Figure 6.**
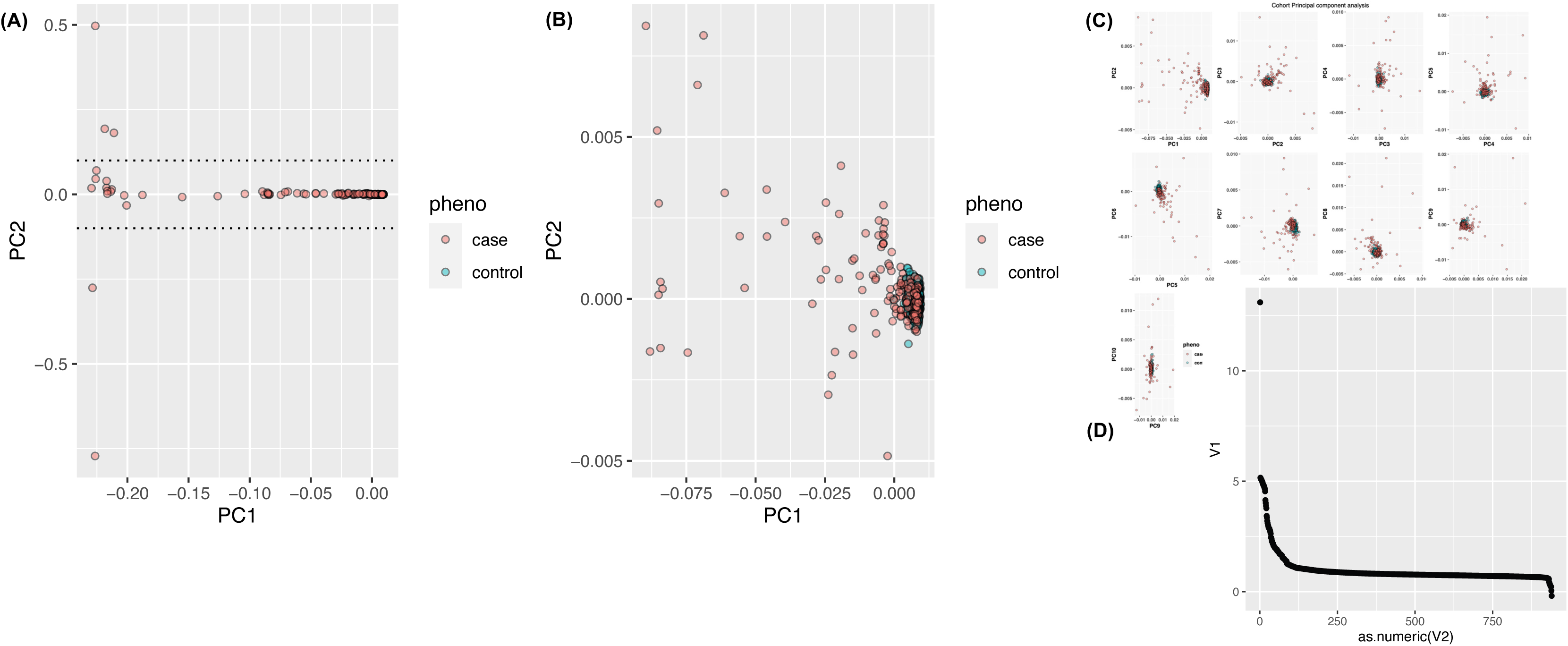
PCA of cohort. Principle component analysis was performed and four samples were dropped from subsequent analysis. PCs were included in statistical models to control for population stratification.

**Figure 7.**
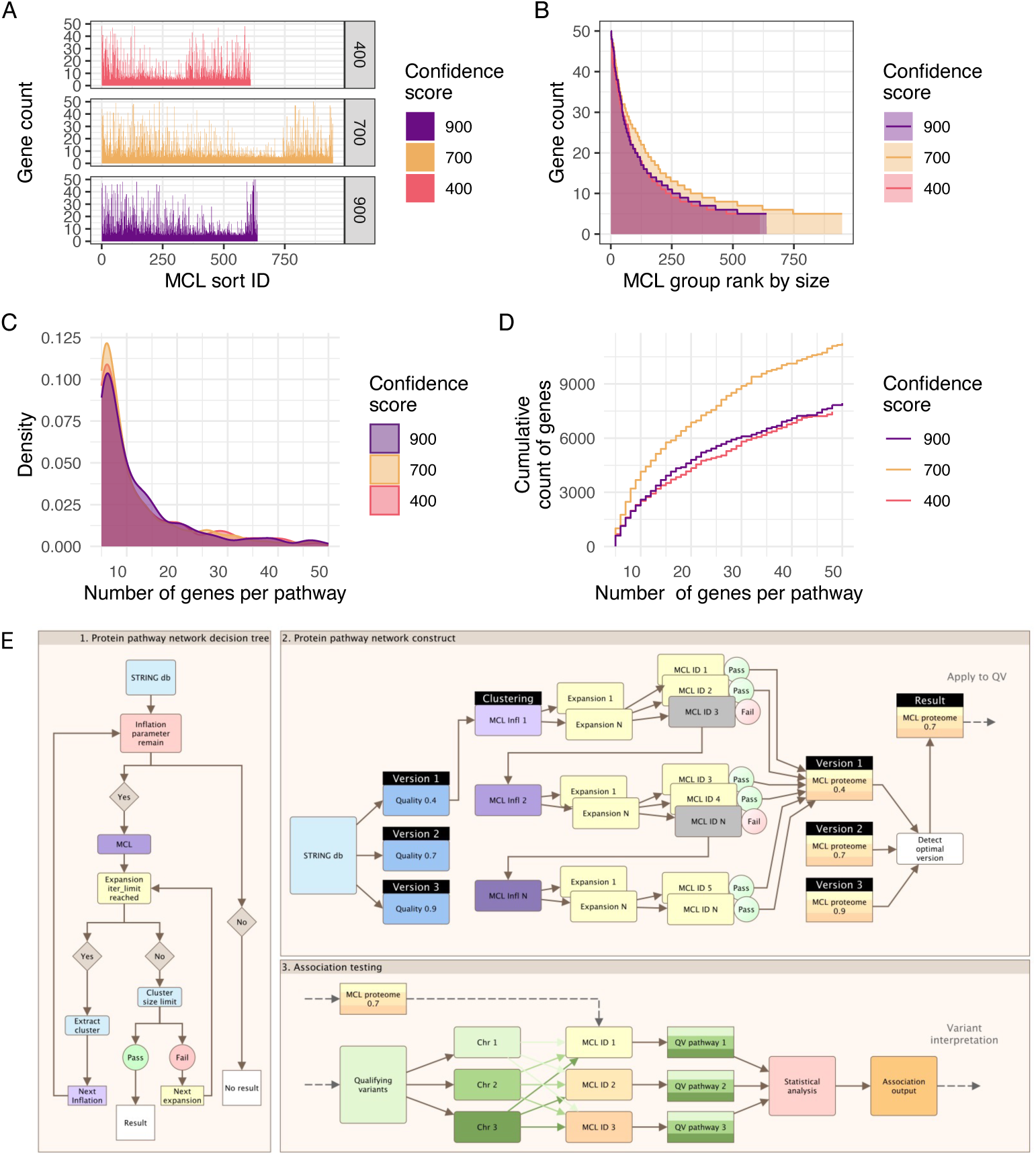
ProteoMCLustR pathway distributions and algorithm implementation. The figures A-D show the summary of genes per pathway distributions after running ProteoMCLustR on the STRINGdb human (9606) v11.5 data for whole genome protein-protein interactions. All three evidence score thresholds (400, 700, 900) are shown. Clustering was optimised to achieve pathway sizes between five and fifty proteins. Markov cluster algorithm (MCL) IDs were used to label protein pathways. (A) The count of genes per pathway sorted by MCL ID order, (B) the gene count ranked by pathway size, (C) the density of genes per pathway, (D) cumulative count distribution of number of genes per pathway. We provide this dataset as the default for ProteoMCLustR repository and these plots are provided as the default “ppi user ouput”. Illustration (E) shows the algorithmic implementation of ProteoMCLustR, optimised for high performance computing, which runs MCL parameters on the input PPI dataset to determine best-fit protein pathways. ProteoMCLustR was automated to use Kruskal-Wallis testing however no significant differences were found in protein counts across confidence score groups. Thus, by default the largest dataset was selected to maximize protein coverage for comprehensive analysis. That is, score 700 provided a 11257 genes qualifying into pathways compared to scores 400 and 900 with counts of 7456 and 7932, respectively.

**Figure 8.**
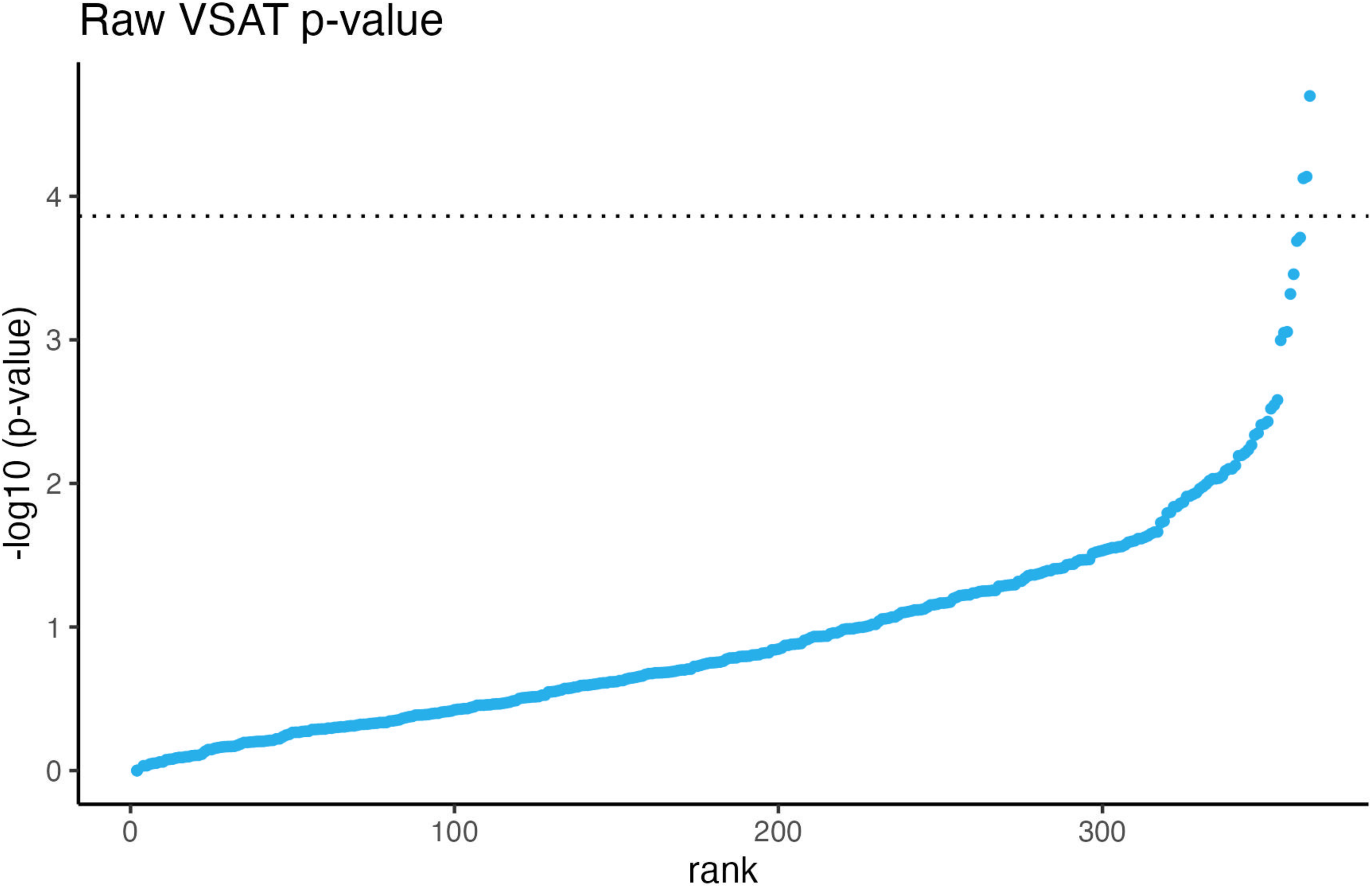
Raw VSAT p-values as used in the Archipelago method. Raw VSAT p-values have no natural x- axis position and must be ranked on association strength or some arbitrary ranking. The same square allelic test. figure 4 (blue) as those shown here without the use of the Archipelago method.

**Figure 9.**
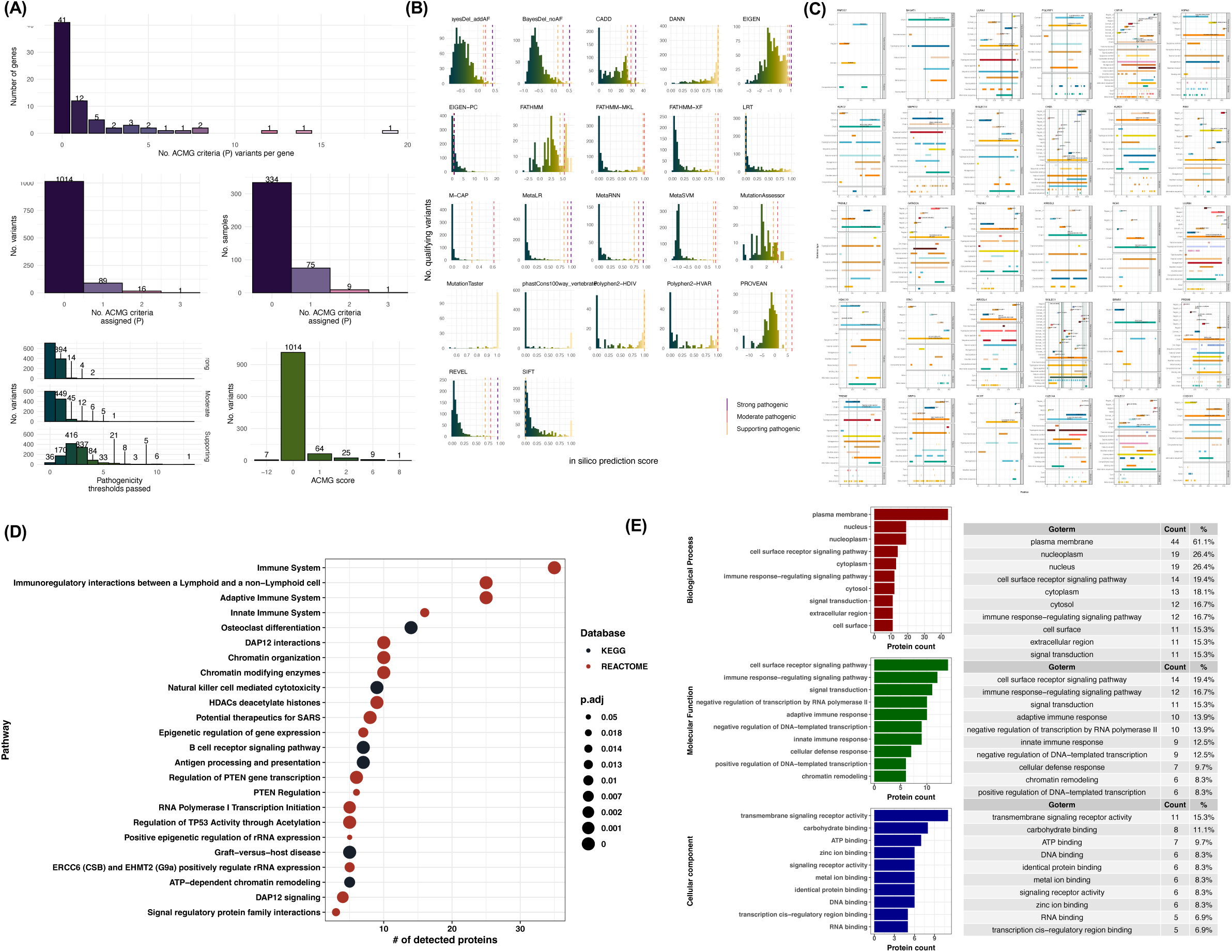
Expanded ACMGuru interpretation of pathogenicity in sepsis-associated protein pathways. ACMGuru gene illustrate visualised the output for ACMGuru post PPI containing the enriched pathways 22, 586, 836. (A) ACMGuru evidence interpretation for protein pathway significantly associated with sepsis: The number of genes versus the number of criteria used for interpreting pathogenicity evidence; the number of unique variants assigned versus the number criteria assigning pathogenicity; the number of samples with unique variants per criteria; additional variant pathogenicity threshold distribution based on reference sources for strong, moderate, and supporting evidence as seen in (B); final determination of ACMG/AMP score by ACMGuru. (B) Distribution of variants for each of the prediction engines with reference sources for strong, moderate, and supporting evidence shown by vertical bars. (C) Gene structure evidence plots and protein structures. Variants are illustrated by vertical bars, with density indicating ACMG scoring. Splicing variants are not present as amino acid positions and are therefore omitted. Genes are only shown which had some evidence of variant effect for interpretation. (D) Combined KEGG, Reactome, and Gene ontology interpretation of the genes reported in the enriched protein pathways with ACMGuru uniprotR.

**Figure 10.**
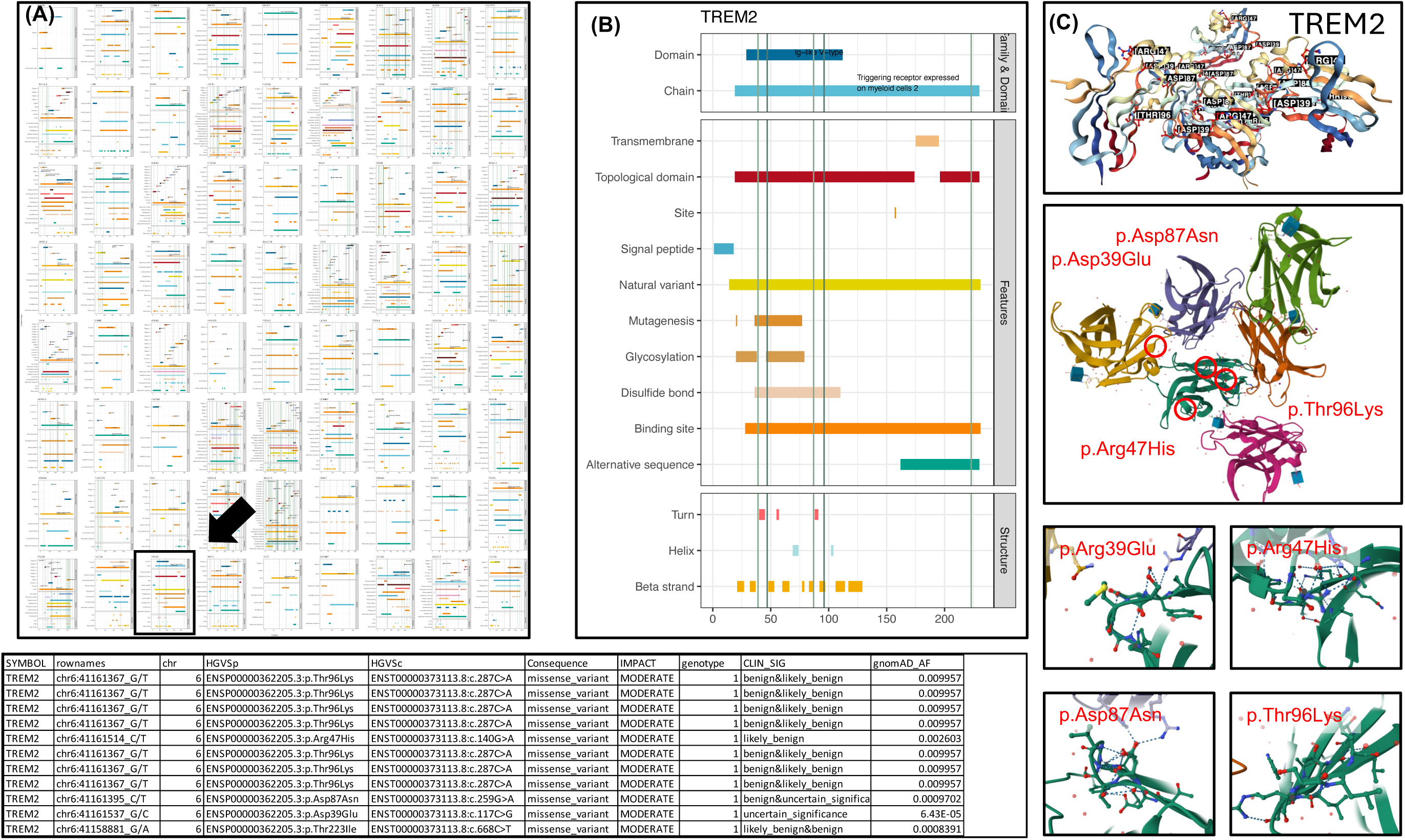
Expanded example ACMGuru interpretation of pathogenicity in an enriched pathway gene. ACMGuru evidence interpretation for a single example gene with variants considered as candidate pathogenic for sepsis. We selected this gene arbitrarily based on our background familiarity with its function to assess the automated interpretation. (A) Gene illustrations showing variants considered pathogenic or likely pathogenic. An example of several variants in TREM2 are highlighted. (B) Gene structure evidence illustration for the highlighted gene. Vertical bars represent the variant position. (C) Automatic protein structure retrieval by ACMGuru AutoDestructR shows all variants affecting this protein, used in the interpretation of mechanism, revealing how variants may influence critical functional domains. Notable affected features include disulfide bonds, glycosylation sites, and the Ig-like V-type domain, key to TREM2’s function in microglial activation and phagocytosis. (Automatically retrieved structure data source: PDB Entry: 5UD7 - Crystal Structure of Wild-Type Ig-like Domain of TREM2. DOI: https://doi.org/10.2210/pdb5ud7/pdb. Primary publication DOI: https://doi.org/10.1074/jbc.RA118.002352).

**Figure 11.**
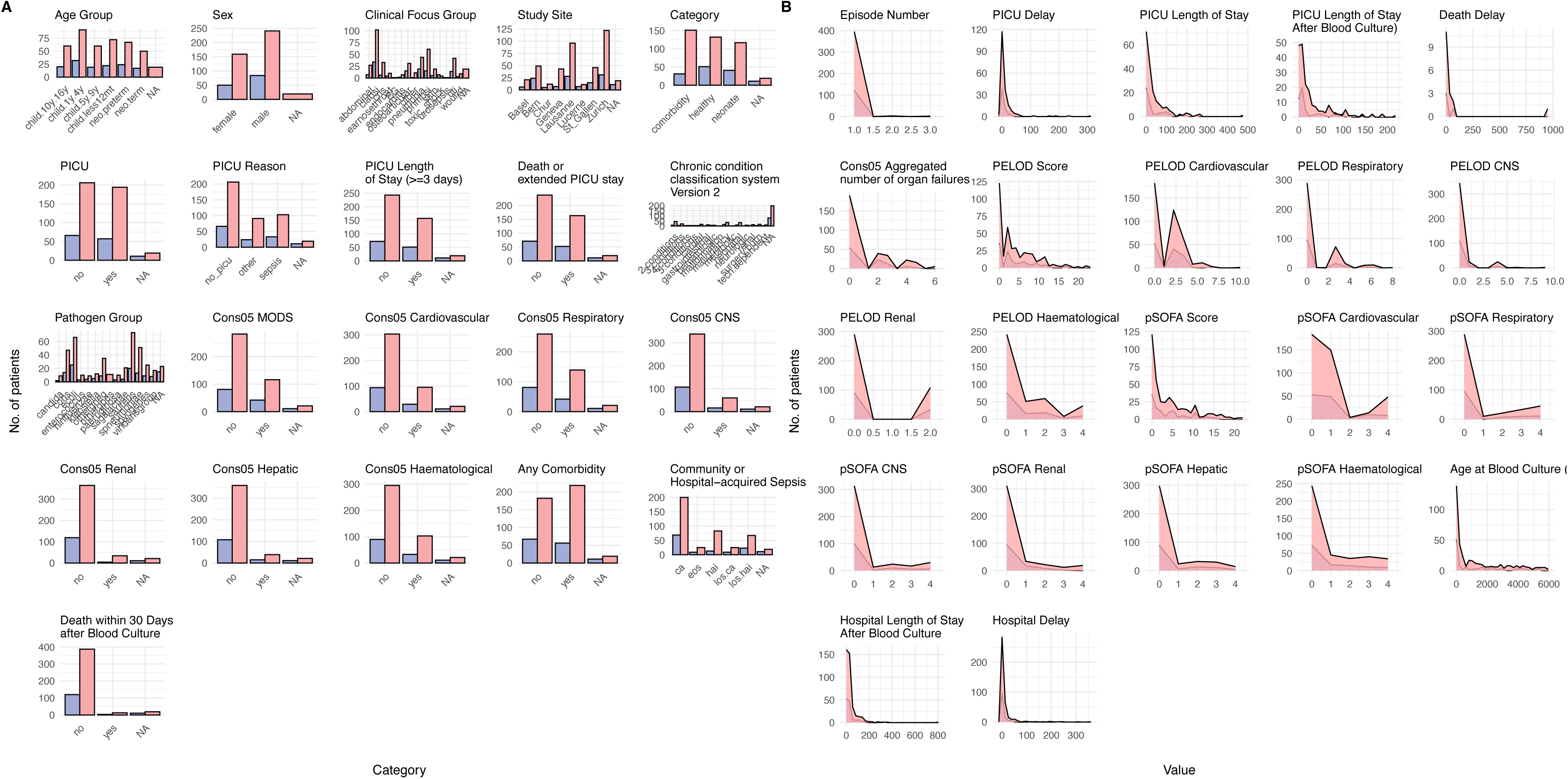
Cohort clinical characteristics separated into carriers of variants in the VSAT enriched pathway versus those whom have no variants in this pathway. No global correlation was expected since each individual is expected to only require one damaging variant within the share pathway.

**Figure 12.**
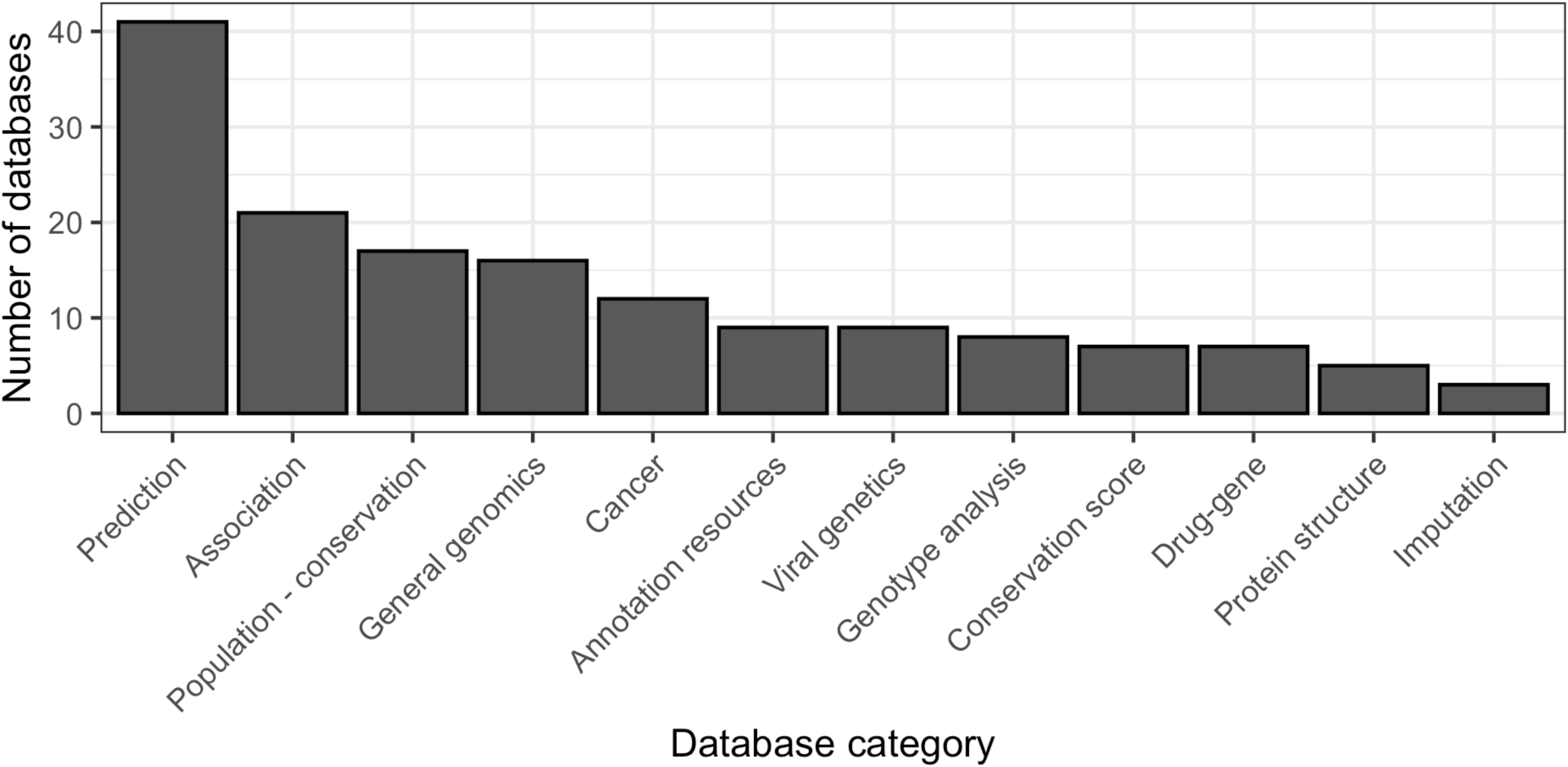
Annotation database categories. Summary of database categories used for variant annotation.

**Figure 13.**
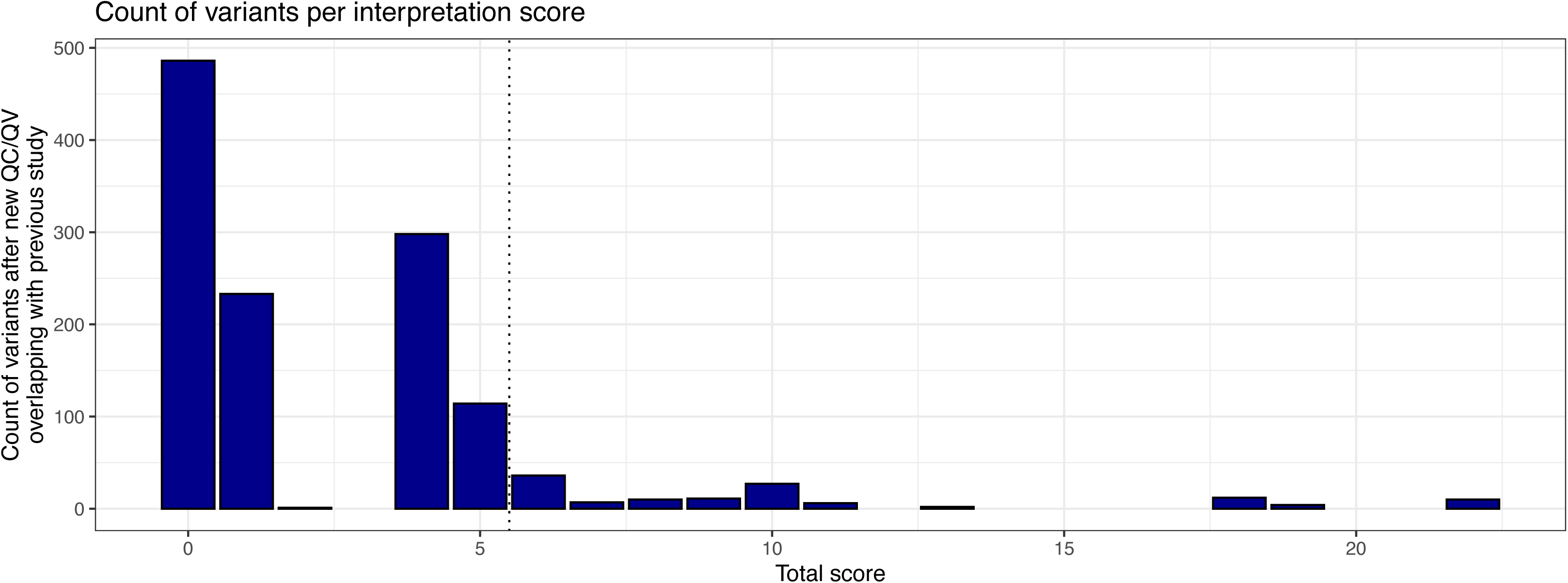
Validation of previous study. A subset of patients (176) were previously analysed in Borghesi et al. (2020), which investigated rare variants in primary immunodeficiency genes in children with sepsis. We screened the new cohort for variants of unknown significance to see if any new candidates were classified. We found that variants were now annotated more accurately with updated databases using standardize scoring criteria. Scores are artificially inflated since we had to remove filters like allele frequency to retain variants during processing.

## 5 Supplemental Tables

**Table 1 Annotation table of data sources used for variant interpretation and filtering.**

Variant annotation is a critical step in clinical genetics. Popular tools for applying annotation data to VCF format genetic data include: NIRVANA https://illumina.github.io/NirvanaDocumentation/, ANNOVAR https://annovar.openbioinformatics.org/en/latest/ Variant Effect Predictor (VEP) http://www.ensembl.org/info/docs/tools/vep/index.html. Additionally, these tools require a set of data sources containing the annotation information which will be applied to each variant. However, each tool generally provides a ready-to-go database that can be utilised. This table represents that databases that have been used on our annotation process.

**Table 2 Cleaned IUIS IEI table.**

This table is a modified version of that released by the International Union of Immunological Societies (IUIS) Inborn Errors of Immunity Committee (IEI) (https://iuis.org),(Tangye et al., 2022): <u>doi.org/10.1007/s10875-022-01289-3</u>. The modification involved cleaning all fields, converting to consistent nomenclature, splitting joined cells, and other miscellaneous tasks to facilitate tabularisation for automation. Additional hyperlinks to external websites have been added along with cleaning and standardisation for filtering “Inheritance” terms. To directly download the IUIS IEI original xlsx table from Springer see: <u>Supplementary file2 (XLSX 93 kb)</u> (https://static-content.springer.com/esm/art%3A10.1007%2Fs10875-022-01289-3/MediaObjects/10875_2022_1289_MOESM2_ESM.xlsx).

**Table 3 Cleaned ACMGuru criteria.**

This table contained the classification criteria used by ACMGuru. Extensive annotation is applied during our genomics analysis. Interpretation of genetic determinants of disease is based on many evidence sources. One important source of interpretation comes from the Standards and guidelines for the interpretation of sequence variants: a joint consensus recommendation of the American College of Medical Genetics and Genomics and the Association for Molecular Pathology (Richards et al., 2015), full text at doi:10.1038/gim.2015.30. The following tables are provided as they appear in the initial steps of our filtering protocol for the addition of ACMG-standardised labels to candidate causal variants.

**Table 4 Cleaned ACMGuru criteria caveats.**

This table contained the classification criteria caveats used by ACMGuru.

**Table 5 ACMG point system for the interpretation of evidence.**

Individual point categories for the naturally scaled point system from Tavtigian et al. (2020) used in combination with the ACMG/AMP variant classification guidelines.

**Table 6 ACMG scoring system for the tally of interpretation of evidence**

Candidate pathogenicity classification thresholds for the naturally scaled point system from Tavtigian et al. (2020) used in combination with the ACMG/AMP variant classification guidelines.

**Table 7 varsome calibrated insilico thresholds used for PP3 criteria.**

This table outlines the calibration of in-silico tools used for classifying missense variant pathogenicity based on the evidence-based approach by VarSome, Saphetor SA, in alignment with ClinGen recommendations. This table presents the calibration outcomes of more than 30 computational predictors against ClinVar data, detailing their accuracy, sensitivity, and specificity. For ACMG criteria PP3 and BP4, evidence thresholds for multiple lines of computational support are delineated, triggering different strength levels of pathogenicity or benign predictions. The final ACMG PP3 score is only assigned when variants meet or exceed the strong, moderate, or supporting pathogenic evidence thresholds, ensuring better reliability. Data sources include GO, KEGG, Reactome, Biocarta, BioCyc, and several experimental databases.

**Table 8 ACMGuru gene/protein function get discussion**

This table was generated using the ACMGuru ‘get discussion’, which queries pre-compiled RDS files (TaxaObj, GeneOntologyObj, and ProteinFunction) to retrieve biological function descriptions for each gene/protein in an enriched VSAT result. The process ensures an automated and unbiased extraction of relevant annotations, providing summary insights into the molecular functions and roles of each gene/protein.

**Table 9 Variants present in significantly enrich PPI pathway associated with sepsis**

This table contains the major genetic feature annotations for all variants that were present in the protein-protein interaction (PPI) pathway which was significantly enriched for the association with sepsis.

**Table 10.**
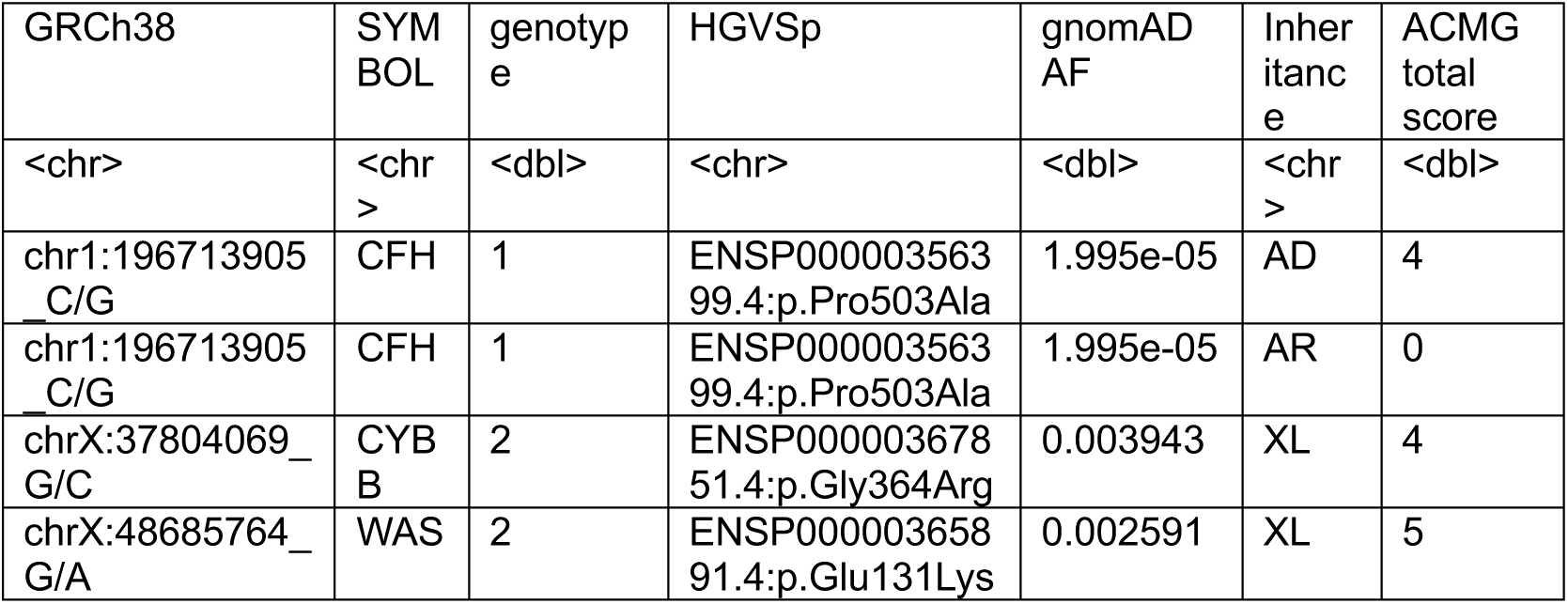
Candidate pathogenic variants previously reported in Borghesi 2020 as re-identified in our study.

